# The Gut Microbiome Strongly Mediates the impact of Lifestyle combined variables on Cardiometabolic Phenotypes

**DOI:** 10.1101/2024.01.12.24301195

**Authors:** Solia Adriouch, Eugeni Belda, Timothy D Swartz, Sofia Forslund, Tiphaine Le Roy, Edi Prifti, Judith Aron-Wisnewsky, Rima Chakaroun, Trine Nielsen, Petros Andrikopoulos, Kanta Chechi, Francesc Puig-Castellví, Philippe Froguel, Bridget Holmes, Rohia Alili, Fabrizio Andreelli, Hedi Soula, Joe-Elie Salem, Gwen Falony, Sara Vieira-Silva, Gianluca Galazzo, MetaCardis Consortium, Jeroen Raes, Peer Bork, Michael Stumvoll, Oluf Pedersen, S. Dusko Ehrlich, Marc-Emmanuel Dumas, Jean-Michel Oppert, Maria Carlota Dao, Jean-Daniel Zucker, Karine Clément

## Abstract

Individual lifestyle factors moderately impact the gut microbiome and host biology. This study explores whether their combined influence significantly alters the gut microbiome and determines the mediating role of the gut microbiome in the links between lifestyle and phenomes. Analyzing 1,643 individuals from the Metacardis European study, we created a non-exhaustive composite lifestyle score (QASD score) incorporating diet quality and diversity, physical activity and smoking. This score shows higher explanatory power for microbiome composition variation compared to individual lifestyle variables. It positively associates with microbiome gene richness, butyrate-producing bacteria, and serum metabolites like Hippurate linked metabolic health. It inversely associates with *Clostridium bolteae* and *Ruminococcus gnavus,* serum branched-chain amino acids and dipeptides observed in chronic diseases. Causal inference analyses found 135 cases where the microbiome mediates >20% of QASD score effects on host metabolome. Microbiome gene richness also emerged as a strong mediator in the QASD score’s impact on markers of host glucose metabolism (27.3% of the effect on HOMA-IR), despite bidirectional associations between the microbiome and clinical phenotypes. This study emphasizes the importance of combining lifestyle factors to understand their collective contribution to the gut microbiota and the mediating effects of the gut microbiome on the impact of lifestyle on host metabolic phenotypes and metabolomic profiles.

## Introduction

Various conditions underpinned by chronic systemic inflammation, from type 2 diabetes and obesity^1–3^ to cardiovascular diseases ^4^ have been associated with gut microbiome alterations. In these diseases, an imbalance in microbial populations (*i.e.*, dysbiosis), is characterized by decreased microbial gene richness and diversity with significant changes in composition and function of the gut microbiome ^5^.

Reports regularly show the importance of a wide array of lifestyle factors in the determination and aggravation of these diseases and suggest a contributing role of the gut microbiome^6,7^. Beyond associations, there is a need to identify microbiome mediators of lifestyle patterns in disease pathophysiology. Amongst lifestyle factors, dietary patterns, physical activity and sedentary behavior, smoking are indeed well-known modifiable factors that can be targeted to improve cardiometabolic health directly or via substantial modifications of the microbiome^8,9^.

Dietary habits can be evaluated through the assessment of the intake of foods and nutrients with protective or deleterious effects on health^10^. Dietary scores have also been proposed to profile population dietary patterns in link to health conditions^10^. Among popular scores, the use of the alternative healthy eating index (aHEI) or the Dietary Approaches to Stop Hypertension (DASH) score for example suggests that better food quality is associated with a reduction in cardiovascular risk^11^. Another score, the alternative Dietary Inflammatory Index (aDII)^12^ also intends to capture the inflammatory potential of the diet. However, only few reports have systematically evaluated the interplay between these score-evaluated dietary patterns and the composition of the microbiome as well as the potential impact on cardio-metabolic eating ^13,14^.

Moreover, eating a diverse diet is generally accepted as a recommendation for promoting nutrient adequacy^15^, but dietary diversity (e.g. defined as a number of different food groups) is not necessarily a proxy of quality^16^. The commonly used dietary diversity scores rarely account for the distribution of food item abundance within each food group considered (in that case named as dietary variety)^16^. Interestingly, the methods to compute microbiome diversity indices can be used to assess diet diversity and variety, thus providing tool to examine relationship with the gut microbiome^17^ and subjects phenotypes. In fact, only a limited number of studies have assessed the relationship between dietary diversity, regardless of dietary quality, and gut microbiome and metabolic health in population with different disease severity ^7,18,19^. Studies have mostly focused on healthy adults or elderly population and did not investigate the associations between dietary patterns and the gut microbiome in the context of cardiometabolic diseases. A previous report found a positive relationship between diet diversity and longitudinal microbial stability in 34 healthy individuals, but no association with microbiome richness^18^. In the PREDICT 1 study, authors showed that the fecal microbiome diversity was associated with habitual dietary intake and diversity^7^. Another study used a score akin to the Simpson index in 445 elderly subjects and described an association between a more diverse diet and an even distribution of bacterial groups^19^.

Strikingly, if these indexes evaluate the healthfulness of food, they do not incorporate other lifestyle factors necessary for effective health promotion recommendations. For instance, the report from the Global Burden of Disease Study (2017) clearly indicates together, with the inadequate fruit and vegetable intake, the harmful nature of alcohol and smoking use, and insufficient physical activity for health^20^. Levels of alcohol consumption has been show to differentially impact gut microbiome in human microbiome studies^7,21,22^. Physical activity^23^ or smoking status^24^ have also been shown to be associated with microbiome composition. Actually, different physical activity or sedentary indexes related to metabolic health can also be used in this context. However, no study has systematically examined the combined effect of these physical activity indexes or smoking status with dietary indexes^25^, in connection with the gut microbiome and metabolic health ^26,27^.

We here evaluated the relationships between a series of dietary components (including alcohol) and indexes (related to diet quality, variety, diversity, and inflammatory potential), physical activity and sedentarity indexes, and smoking status and the gut microbial gene richness and composition in a large-scale study involving 1,643 participants from the European MetaCardis project with a broad spectrum of cardiometabolic phenotypes. We created a combined lifestyle score (called QASD), encompassing diet quality and diversity, physical activity and smoking status, and examined its relationship with metabolic health, serum and urinary metabolome, and fecal microbiome. Most importantly, to gain a deeper understanding of how this new composite lifestyle score interacts with host cardiometabolic health via potential microbiome contribution, we carried out causal inference (e.g., mediation) analyses, providing valuable perspectives on these intricate relationships.

## Results

### Lifestyle and bioclinical characterization of Europeans with and without cardiometabolic dysfunctions

The 1643 studied individuals of the MetaCardis population include a healthy group (n=305) and different pathology groups as follows: individuals with metabolic syndrome (n=220), type 2 diabetes (n=477), obesity (BMI ≥30, n=222), severe obesity (BMI ≥35 n=112), and cardiovascular diseases (n=307) (Supplemental Table 1). Healthy individuals and individuals with obesity were younger (p<.0001) and more often female (p<.0001) than in the other groups. Pharmacological treatments were used and have been identified as factors sometimes masking the disease related signatures of the gut microbiome^28,29^. Thus, medication such as five-year retrospective exposure of antibiotics (total number of courses), metformin, statin and protein pump Inhibitors (PPI) usage were included as covariates in further analysis. Supplementary and sensitive dietary analyses were adjusted on energy intake in kcal and aHEI score for dietary analysis and on BMI or MetaCardis pathology group for metagenomic analyses, e.g., see Methods section and legends for the adjustment strategies used.

Statistical observations acquired in individuals with overweight and obesity from the MetaCardis study were replicated in the GutInside cohort composed of French adults with overweight and obesity (n=433) that were recruited in different French regions between September 2018 and January 2020^30^. Only baseline data were used in this dietary intervention program. Subjects’ phenotypes are presented in Supplemental Table 2. Importantly, both GutInside and MetaCardis projects applied similar lifestyle questionnaires: e.g. Food Frequency Questionnaires (FFQ) for habitual food intake^31^, the Recent Physical Activity Questionnaire (RPAQ) for physical activity^32^ and gathered smoking-related information, including past habits and current cigarette consumption. Shotgun metagenomic sequencing was available in both cohorts.

### Lifestyle variables associate with gut microbiome gene richness

We examined the relationships between Gut Microbiome Gene Richness (GMGR, for number of microbial genes from Integrated Gene Catalog (IGC) detected in metagenomic samples) and lifestyle variables including nutrients (n=72 variables), food groups (n=68 variables), physical activity variables (n=37) and smoke habits (N=3, as qualitative variables). Out of 177 quantitative variables analyzed, 43 (24.29%) of them were significantly associated with GMGR (FDR<0.05; partial Spearman correlation analyses ; Figure 1A). The adjusted effect sizes were modest (Spearman Rho ranging from -0.12 to 0.13). Lifestyle variables consistently and positively associated with GMGR were for food groups: intake of alcoholic beverages (e.g., wine and beer), tea and coffee, nuts and fish. For nutrients, we found choline, vitamins A/E, B7/B8 or biotin, D, mono-unsaturated fatty acids and omega 6 fatty acids. We also observed positive associations between GMGR and intake of total polyphenols, chalcones (found in beer), isoflavonoids (soja, onions, wine and tea) and hydroxycinnamic acids (found in coffee). These nutrient- based results are in agreement with observations at the food group level. Moreover, these observations are in line with reported association between polyphenols and gut microbiome markers and overall health^33,34^. Our analysis also revealed a negative association with trans fatty acids, which have been previously linked to bacteriostatic additives or diet-induced inflammation^35^, and with the intake of simple sugars at nutrient level (Figure 1A). At food group level, we observed a negative association between total amount of starches and GMGR (rho=-0.08), mostly driven by the intake of simple sugars. This food group shows the strongest negative association with GMGR (rho=-0.12 ; Figure 1A). This negative association was reproduced in stratified analyses within individual clinical subgroups. On the other hand, a positive association between the intake of fruits and GMGR was found in the healthy group (Supplemental Figure 1A; rho=0.22, FDR<0.05) in line with previous results^36^. Looking at overall dietary scores (e.g., aHEI, DASH, and the inflammatory score aDII), the aHEI score, that reflects the overall diet quality, showed a significant positive association with GMGR, but no significant association was found with DASH or aDII (Figure 1B).

**Figure 1.**
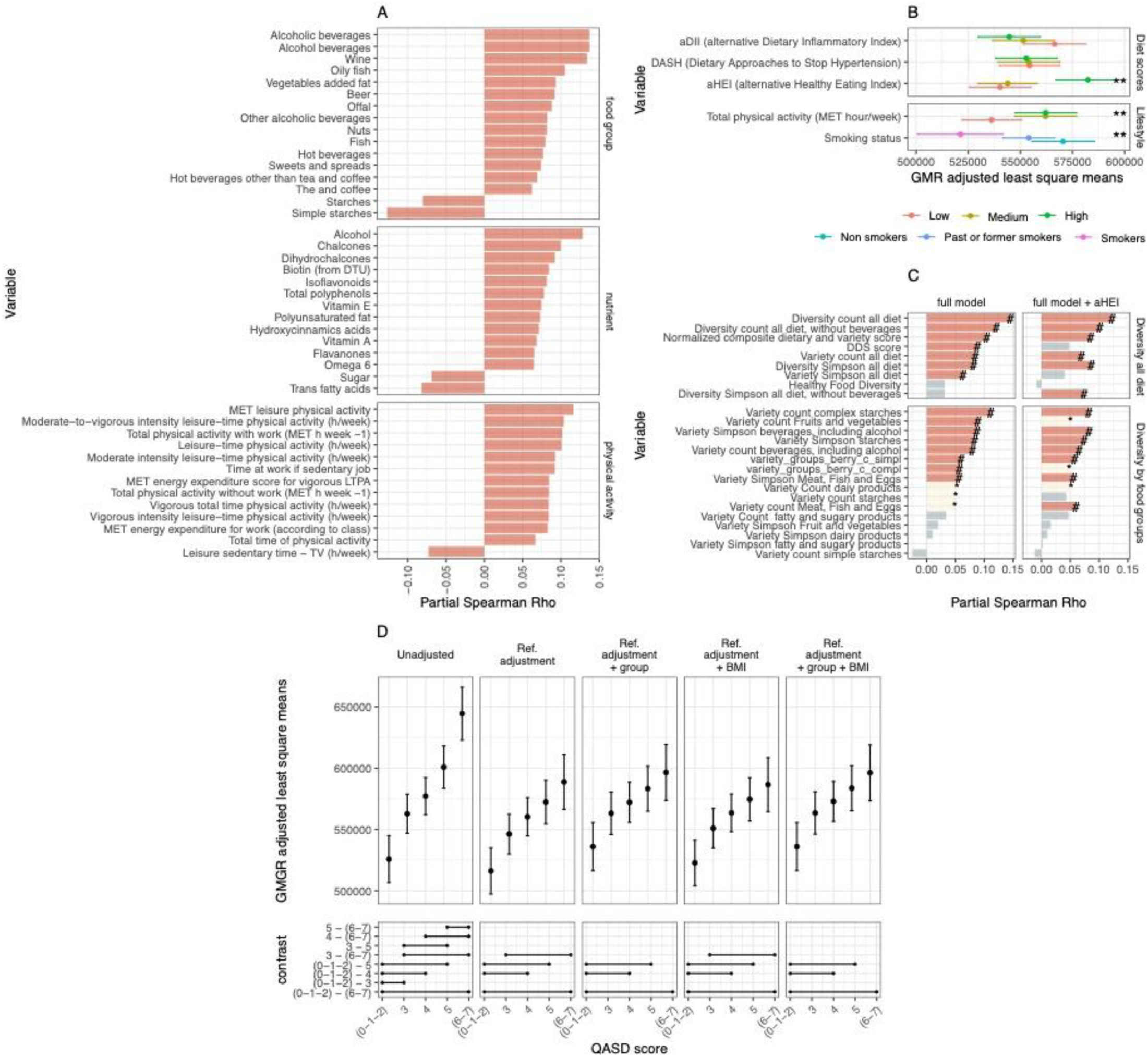
alternative Dietary and physical activity factors, diet variety and diversity scores and a composite lifestyle score associated with Gut Microbiota Gene Richness (GMGR) in MetaCardis population. (A) Barplot of 43 food groups, nutrients and physical activity variables significantly associated with GMGR (FDR<0.05; partial Spearman’s rank coefficient correlation with GMGR, adjusted on age, center of recruitment, energy intake in kcal, antibiotics treatments (number of treatments in past five years), metformin (yes/no), statin (yes/no) and PPI (yes/no) medications. (B) Dietary scores (aHEI, DASH, aDII) and lifestyle factors (Total physical activity and Smoking status) associated with GMGR. For diet scores and total physical activity, the variables were divided in tertiles (low/medium/high), and estimated marginal means of GMR across levels of the variables derived from linear regression models adjusted on age, center of recruitment, energy intake in kcal, antibiotics treatments (number of treatments in past five years), metformin (yes or no), statin (yes or no) and PPI (yes/no) medications are represented (**=FDR<0.05; ANOVA tests on linear regression models). (C) Associations of 25 Dietary diversity and variety scores with GMGR (*: p-value<0.05; #: FDR<0.05; partial Spearman’s rank correlation). Full adjusted model corresponds to partial correlations adjusted on age, center of recruitment, energy intake in kcal, antibiotics treatments (number of treatments in past five years), metformin (yes or no), statin (yes or no) and PPI (yes/no) medications, whereas full adjusted model + aHEI are partial correlations additionally adjusted by alternative Healthy Eating Index in order to evaluate if the association are independent of diet quality. Scores refer on all diet (top columns) or within food groups (bottom columns). (D) Estimated Marginal Means of GMGR across levels of the QASD score in total MetaCardis study unadjusted and adjusted by different covariates based on linear regression analyses. Reference adjustment corresponds to models adjusted by age, center of recruitment, energy intake in kcal, antibiotics treatments (number of treatments in past five years), metformin (yes or no), statin (yes or no) and PPI (yes/no) medications. Contrast panel connects pairwise levels of the QASD score with significant differences in estimated marginal means (p-value<0.05; post-hoc correction for multiple comparisons with Tukey method)

Furthermore, we examined other lifestyle variables. Increased in physical activity during work and leisure time (representing total physical activity) was positively linked to a higher GMGR, whereas the time spent in sedentary activities in front of television screens (representing total sedentary behavior) exhibited a negative association with GMGR (Figure 1A). Regarding categorical variable, smoking status associated with GMGR, with smokers displaying an inverse association with GMGR (Figure 1B). These associations were robust to additional adjustments by clinical group and BMI status (Supplemental Figures 1 C, D).

These first analyses revealed statistical links between several lifestyle variables and GMGR in agreement with investigations performed in general populations^7,37^. Hence, the disparity in the ratio of positive and negative associations with GMGR, coupled with the moderate effect sizes observed for individual variables, prompted us to conduct a detailed exploration of the relationships between GMGR and dietary diversity (measured as the number of food groups) and dietary variety scores (measured as the number of different foods within the same group), as well as with a combined lifestyle score.

### Dietary diversity and variety are associated with microbiome gene richness regardless of dietary quality

We calculated 25 scores of dietary variety and diversity and examined their statistical relationships with GMGR. These scores take into account diet as a whole (identified by the term “diversity all diet”), specific food groups (identified by “variety”), simple counts in food consumption within groups (identified by “count”), the evenness of consumption within groups (identified by “Simpson”) (See Methods, Supplementary Table S4 for full details of scores calculation and sensitivity analyses). We calculated the normalized composite dietary and variety score of the diet^38,39^, a score that combines diversity and variety and the Healthy Food Diversity index (HFD) which evaluates the diversity and food quality ^40^. Fifteen of these scores were positively associated with GMGR (Figure 1C, Partial Spearman rho >0, FDR<0.05, full adjusted model), including 7 of the 9 scores describing diet diversity. Interestingly, 11 of these associations remained significant after adjustment for the dietary quality as assessed by the aHEI score, including 2 additional scores (« Diversity Simpson all diet, without beverages » and « the Variety count Meat, Fish and Eggs ») where the association with GMGR were even strengthened (Figure 1C, full adjusted model + aHEI, see methods).

Stratifying these scores by food groups, we identified that the variety count of complex starches was the score with the strongest positive association with GMGR, followed by the variety of “fruits and vegetables” (defined by both count and Simpson), the variety of “beverages, including alcohol” (« count and Simpson »; rich in polyphenols) and the variety Simpson of “meat, fish, eggs” (Figure 1C). On the opposite side, we observed that the Variety count of simple starches was a score with a negative association with GMGR, even if non-significant at statistical level. This is in line with the negative association observed at the nutrient and food group level with the amount of sugar and simple starches described in Figure 1A.

Importantly, the observed positive associations extended to the diversity of products that are not considered to be healthy food items like variety of “beverages, including alcohol”. The score “variety of all the diet” was the score most strongly associated with GMGR. Contrary to previous observations^7^, the HFD score was not associated with GMGR in the MetaCardis population considering the entire cohort and individual clinical subgroups (Figure 1C, Supplemental Figure 1B).

Overall, food diversity and variety within specific food groups show positive associations with GMGR, irrespective of their healthfulness, in this population with a high prevalence of metabolic and cardiovascular diseases.

### A newly developed composite lifestyle score is associated with gut microbial gene richness, even when stratified by the severity of cardiometabolic phenotypes

Recognizing that isolated statistical effects do not capture additive effects, interaction, and intricate combinations of lifestyle variables, and considering the relatively modest effect sizes of these individual variables, we hypothesized that a combination of different lifestyle variables could provide a more significant advantage in explaining GMGR variability than individual effects. We thus constructed a composite lifestyle score, named ’QASD’ (acronym for dietary Quality, physical Activity, Smoking, dietary Diversity), by assigning tertile points to diet quality (aHEI, 0/1/2), total physical activity (total physical activity in MET-h.week-1, 0/1/2), and diet diversity excluding beverages (Simpson diversity score for all foods except beverages, 0/1/2). This food diversity score was favored because it showed an independent association with GMGR, separate from aHEI. For smoking status, 0 point for smokers and 1 for non-smokers were attributed. Of note the aHEI comprises the consumption of alcohol that was strongly associated with microbiome richness in univariate analysis.

Using an additive approach, where each element carries equal weight, patients received scores between 0 (indicating the lowest QASD) and 7 (indicating the highest QASD); a higher QASD representing a more favorable lifestyle). Interestingly, we found strong association between the QASD score and Gene Richness in the entire MetaCardis population, that remained significant after several adjustment models (Fid 1D; p-value<0.05, ANOVA test on linear regression models of GMGR by QASD score and additional cofounders; p-value<0.05 on Tukey post-hoc pairwise tests across QASD score levels). We next investigated if and how the interaction between QASD score and GMR would impact on the clinical profile of individuals.

### The QASD score exerts a significant impact on both gut microbiome and metabolic status of individuals

We evaluated statistical interplay between the QASD score, GMGR and individual clinical profiles considering routine phenotype; e.g., Body Mass Index (BMI) and glucose metabolic status (using glycated hemoglobin for glucose control and HOMA-IR, as a surrogate of insulin resistance) by conducting bi-directional mediation analyses under two different hypotheses with the QASD score as the independent variable (Figure 2A). The first hypothesis tests whether the QASD score influences the GMGR through variation in individual’s clinical profiles (as mediators). The second hypothesis proposed that the QASD score impacts the clinical profile, via GMGR variation (as a mediator). We performed this analysis considering the extremes of the QASD score (QASD score 0-1-2-3; n=662 vs. QASD score 5-6-7; n=546) as well as each of the extremes (e.g., “low” QASD or “high” QASD) versus intermediate QASD score level (QASD=4; n=435). Interestingly, this mediation analysis provided support for both hypotheses regarding the extremes of the QASD score and metabolic status (high vs. low, Figure 2B, P-value<0.05 for the total effect, Average Direct Effect (ADE) and Average Causal Mediation Effect (ACME); mediation analyses adjusted by age, gender, country of origin, and intake of Metformin, Statin and PPI).

**Figure 2:**
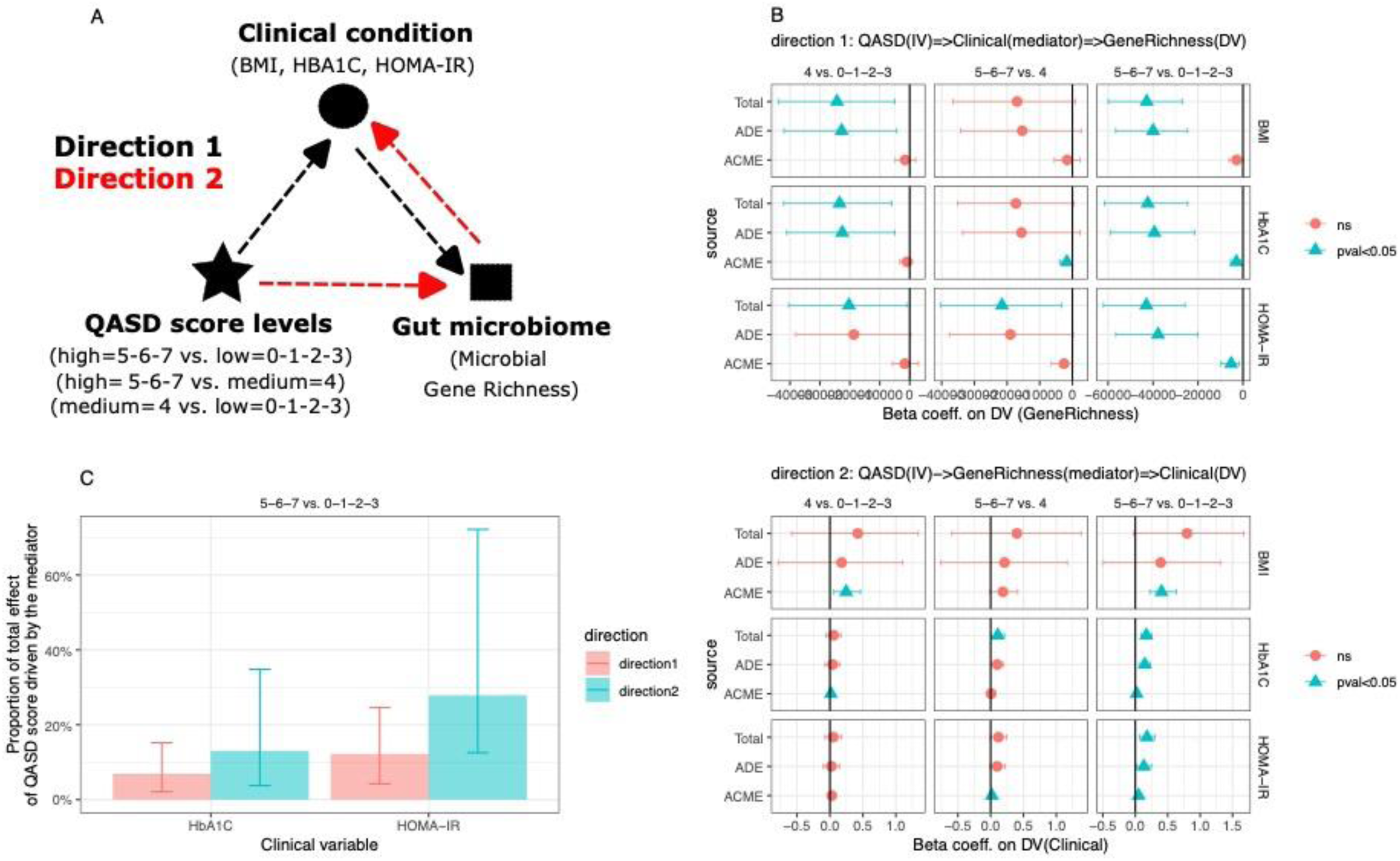
Causal mediation analyses on the impact of QASD score on the gut microbiome and clinical conditions in Metacardis population. (A) Schematic representation of the two-mediation hypothesis tested to evaluate the impact of QASD score (independent variable) on the gut microbiome and clinical profile of individuals. In direction 1 the QASD score would impact gut microbiome (GMGR; dependent variable) mediated by alterations in clinical profile (mediator). In direction2 the QASD score would impact the clinical profile (dependent variable) mediated by alterations in the gut microbiome (GMGR; mediator). BMI, glycated haemoglobin and HOMA-IR have been tested as clinical variables. Pairwise QASD score levels compared were low (0-1-2-3) vs. medium (4), low vs. high(5-6-7), and medium vs. high. Mediation analyses were adjusted by center of recruitment, age, gender and intake of metformin, statin and PPI. (B) Confidence intervals of the beta coefficients corresponding to the total effect (Total), Average Direct Effect (ADE) and Average Causal Mediation Effect (ACME) of the QASD score on the dependent variable in each direction (GMGR in direction 1; clinical variables on direction 2). Color and shape reflect the significativity of the effects in mediation analyses. Only for extreme levels of the QASD score (high vs. low) vs. glycated haemoglobin and HOMA-IR we observed an overall significant mediation in both directions (P-value<0.05 for Total, ADE and ACME) (C) Decomposition of these overall significant mediations as a barplot representing the proportion of the total effect and confidence intervals of the extremes of the QASD score (high vs. low) on the dependent variable (GMGR in direction 1; clinical variable in direction 2) that is driven by the mediator (clinical variable in direction 1; GMGR in direction 2)

In line with hypothesis 1, variations in HOMA-IR and glycated hemoglobin levels explained 12.2% and 6.8%, respectively, of the influence of the QASD score on GMGR (Figure 2C). However, no significant mediation was found using BMI. However, our results also underscore that QASD score significantly affects individuals’ glucose metabolism status via GMGR. We found that a significant proportion of this effect could be attributed to alterations in the gut microbiome, in line with hypothesis 2. This mediation accounted for 27.3% of the QASD score’s effect on HOMA-IR and 13.0% on glycated hemoglobin level.

Overall, this bi-directional mediation analyses reveal that lifestyle (e.g., evaluated by our score as a surrogate) influences gut microbiome gene richness through variations in clinical profiles but reciprocally affects clinical profiles via GMGR variation and in that case with a stronger mediating effect.

### The QASD score explains gut microbiome composition variation even when stratified by enterotype

GMGR represents only a facet of microbiome composition, we further investigated the relationships between the QASD score and compositional aspects of the gut microbiome. We used distance-based redundancy analyses (dbRDA) and observed that 47 lifestyle variables strongly associated with microbiome composition (Figure 3A; FDR<0.01, Supplemental Table S5). The aHEI (adjR2=4.72e- 03, FDR=4.62e-03) and the QASD score (adjR2=3.89e-03, FDR=4.62e-03) showed the highest effect size, followed by the intake of « sweets and spreads » (adjR2=3.56e-03; FDR=4.62e-03), of « hydroxycinnamic acids » (adjR2=2.27e-03, FDR=4.62e-03), “alcohol” (adjR2=2.1e-03, FDR=4.62-03) and of food vitamins like vitamin D (adjR2=2.24e-03, FDR=4.62e-03), vitamin A (adjR2=1.28e-03, FDR=4.62e-03) or biotin (adjR2=1.02e-03, FDR=4.62e-03). Interestingly here, a significant association was also found between the inflammatory potential of the diet and microbiome composition (aDII score; adjR2=1.18e-03, FDR=7.04e-03) (Figure 3A; Supplemental Table S5).

**Figure 3:**
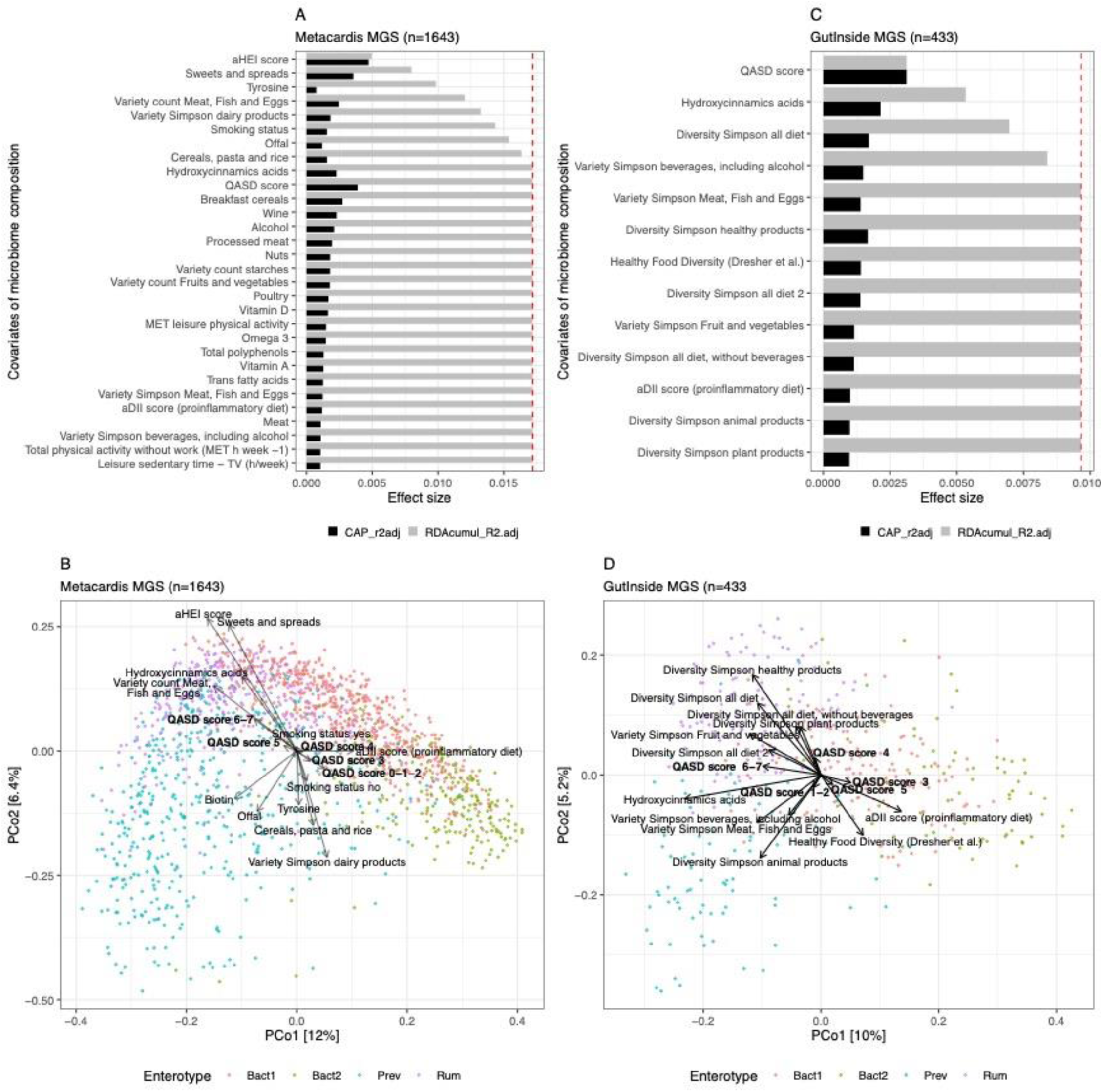
Evaluation of the links between lifestyle on microbiome composition in the Metacardis and GutInside cohorts: (A) 30 of the 47 nutritional variables with highest significant impact over microbiome composition in individuals of the MetaCardis cohort (n=1643; distance-based redundancy analysis, dbRDA; Bray-Curtis dissimilarity from MGS abundance data), either independently (univariate effect sizes in black; FDR<0.01 in dbRDA) or in a multivariate model (cumulative effect sizes in gray, n=9 variables). The cut-off for a significant non-redundant contribution to the multivariate model is represented by the red line (p-value<0.05 in stepwise model building). Full results in Supplemental Table S5. (B) Principal coordinates analysis of inter-individual differences (Bray–Curtis dissimilarity from MGS abundance data) in the microbiome profiles of individuals from the MetaCardis cohort (n = 1643). Arrows represent the effect sizes of a post hoc fit of 9 significant nutritional covariates identified in the multivariate model of panel A together with the QASD score (bold), the proinflammatory diet score and nutritional biotin. (C) 13 Nutritional variables with significant impact over microbiome composition in individuals of the GutInside cohort (n=433; distance-based redundancy analysis, dbRDA; Bray- Curtis dissimilarity from MGS abundance data), either independently (univariate effect sizes in black; p-value<0.05 in dbRDA) or in a multivariate model (cumulative effect sizes in gray, n=5 variables). The cut-off for a significant non-redundant contribution to the multivariate model is represented by the red line (p-value<0.05 in stepwise model building). (D) Principal coordinates analysis of inter-individual differences (Bray–Curtis dissimilarity from MGS abundance data) in the microbiome profiles of individuals from the GutInside cohort (n = 433). Arrows represent the effect sizes of a post hoc fit of 13 nutritional covariates identified in the dbRDA analyses (panel C).

From these 47 lifestyle variables showing significant association with microbiome composition in the univariate dbRDA step, we conducted a step-wise forward model selection by permutation and we obtained 9 variables with a non-redundant explanatory power on microbiome compositional variation. This multivariate model explained 1.72% of compositional variability. These 9 variables included two individual components of the QASD score (e.g., “aHEI score” and “Smoking status”), the intake of “Sweets and spreads”, Tyrosine, Offal, “Cereals, pasta, rice” and “Hydroxycinnamic acids” as well as the “Variety count of Meat, Fish Eggs” and “Variety Simpson dairy products” (Figure 3A).

Moreover, environmental fitting of the 9 lifestyle variables included in the multivariate model into the PCoA ordination landscape generated from Bray-Curtis beta-diversity measures, revealed that aHEI score, the intake of “sweets and spreads” and “hydroxycinnamic acids” and the variety of “meat, fish and eggs” strongly associated to the microbiome compositional space dominated by the *Ruminococcus* enterotype (Rum). This enterotype was previously associated to higher GMGR^41^. The intake of tyrosine, offal, “cereals, pasta and rice” and the variety of dairy products points to opposite direction in the compositional space (Figure 3B). The additional inclusion of the QASD score and the aDII score in the environmental fitting shows that high QASD score levels were also associated with the compositional space dominated by Rum enterotype, whereas the aDII and lower QASD score levels (0-2) were associated to compositional landscape dominated by *Bacteroides* 2 enterotype (Bact2), previously associated to both a lower GMGR and bacterial load, systemic inflammation and obesity severity^30,41^.

Importantly, logistic regression analyses of enterotype status vs. 47 nutritional covariates retrieved in the dbRDA step adjusted by age, gender, BMI center of recruitment, metformin, statin, and protein PPI intake statistically support the relationships between diet variables and a dysbiotic microbiota. We observed a significant decrease in the probability of Bact2 composition with increased levels of the QASD score, the variety of « meat, fish and eggs », offal, biotin, vitamin D but also with and the intake of « cookies and pastries » emphasizing again on the importance of diet diversity and variety (p-value<0.05; Supplemental Figure 2, Supplemental Results). Interestingly, environmental fitting of biotin (vitamin B7/8 calculated from food consumption) on the PCoA ordination landscape pointed towards regions of the compositional space dominated by *Prevotella* enterotype, a microbial composition dominated by bacteria auxotrophic for biotin production^3^ (Figure 3B), which is line with the significant positive association of biotin intake with the probability of Prevotella enterotype in the logistic regression analyses (Supplemental Figure 2, Supplemental Results). Several associations were reproduced in the second independent cohort of 433 individuals with overweight and obesity from the GutInside study^30^, where the QASD score was the variable with the highest and non-redundant explanatory power in microbiome compositional variation (Figure 3C). Environmental fitting showed that high QASD was also associated to *Rum* enterotype, whereas the aDII score was associated to *Bact2* enterotype (Figure 3D).

Extended analyses on Metacardis population with nutritional and bioclinical variables showed that beside corpulence variables (BMI, fat mass), diabetes status or metformin intake, which showed the highest explanatory power, aHEI and the QASD score were the most important lifestyle variables, with an explanatory power higher than covariates like statin intake, lipid and hepatic variables (FDR<0.01, dbRDA analyses; Supplemental Figure 3A). Other components of the QASD score (smoking status, Total physical activity without work (MET h week −1)) were also retained as significant in this extended dbRDA analyses (FDR<001; Supplemental Figure 3A). aHEI and smoking status were also included in a multivariate model of 22 nutritional and clinical variables showing non-redundant explanatory power on microbiome compositional variation, suggesting that QASD score and clinical profiles exerts a non-redundant impact on microbiome (Supplemental Figure 3B). Finally, QASD score association with microbiome composition was robust to cofounding effects in adjusted dbRDA analyses by age, gender, country, BMI, metformin, statin and PP intake (Supplemental Figure 3C).

These observations underscore the robust relationships between the new QASD score and a diversity of nutritional variables and the microbiome composition. We discovered new associations between the QASD score and the diversity of several food items and the lower prevalence of the dysbiotic Bact2 enterotype, that was also replication in an independent population.

### The QASD score associates with metagenomic species and metabolomic signatures

Our next aim was to assess which metagenomic (taxonomic and functional) and metabolomic characteristics (serum, urine) were linked to the QASD score within subjects from the MetaCardis cohort. To discern the potential influence of medication (metformin, statin, and PPI intake) and clinical status on these associations, we applied a post hoc filtering approach utilizing the *metadeconfoundR* R package, as previously developed^29^. Among the various feature examined, the MGS and serum metabolites exhibited the highest number of significant and rigorously deconfounded associations, totaling 194 MGS and 319 metabolites with an FDR < 0.1 under OK_sd status. Notably, the majority of these statistically significant associations surfaced between the extremes of the QASD score (0-1-2-3, low QASD, vs. 5-6-7, high QASD) (Supplemental Figure 4). In contrast, only 0, 6, and 11 features were retained in urine, functional modules (GMM), and mOTUs, respectively (Supplemental Figure 4; detailed results in Supplemental Table S7). Of these, we focused the subsequent analyses on 135 MGS and 272 metabolites with absolute effect sizes (Cliff’s delta) higher than 0.1. At MGS level, higher QASD score levels showed an enrichment in various Firmicutes lineages including *Faecalibacterium prausnitzii* (48 MGS belonging to the *Faecalibacterium* genus and 12 MGS corresponding to different *F. prausnitzii* strains (SL3/3 and L2-6)), *E. eligens*, or multiple MGS of the *Oscillibacter* and *Roseburia* genus, whereas reduced QASD scores were linked to increased levels of *C. bolteae* and *Ruminococcus gnavus* lineages (Figure 4; Supplemental Results and Supplemental Table). Machine learning models, trained for pairwise classification of QASD score levels using these deconfounded MGS signatures, exhibited modest performance on hold-out datasets. The best Area Under the Curve (AUC) values were achieved by random forest (63.7) and ternary Predomics models (55.65) in high vs. low QASD score classification. The inclusion of *C. bolteae* and *R. gnavus* proved important in optimizing the predictive capability of these models for classification between pairwise QASD score levels (see Supplementary Figures 5, 6; Supplemental Results).

**Figure 4:**
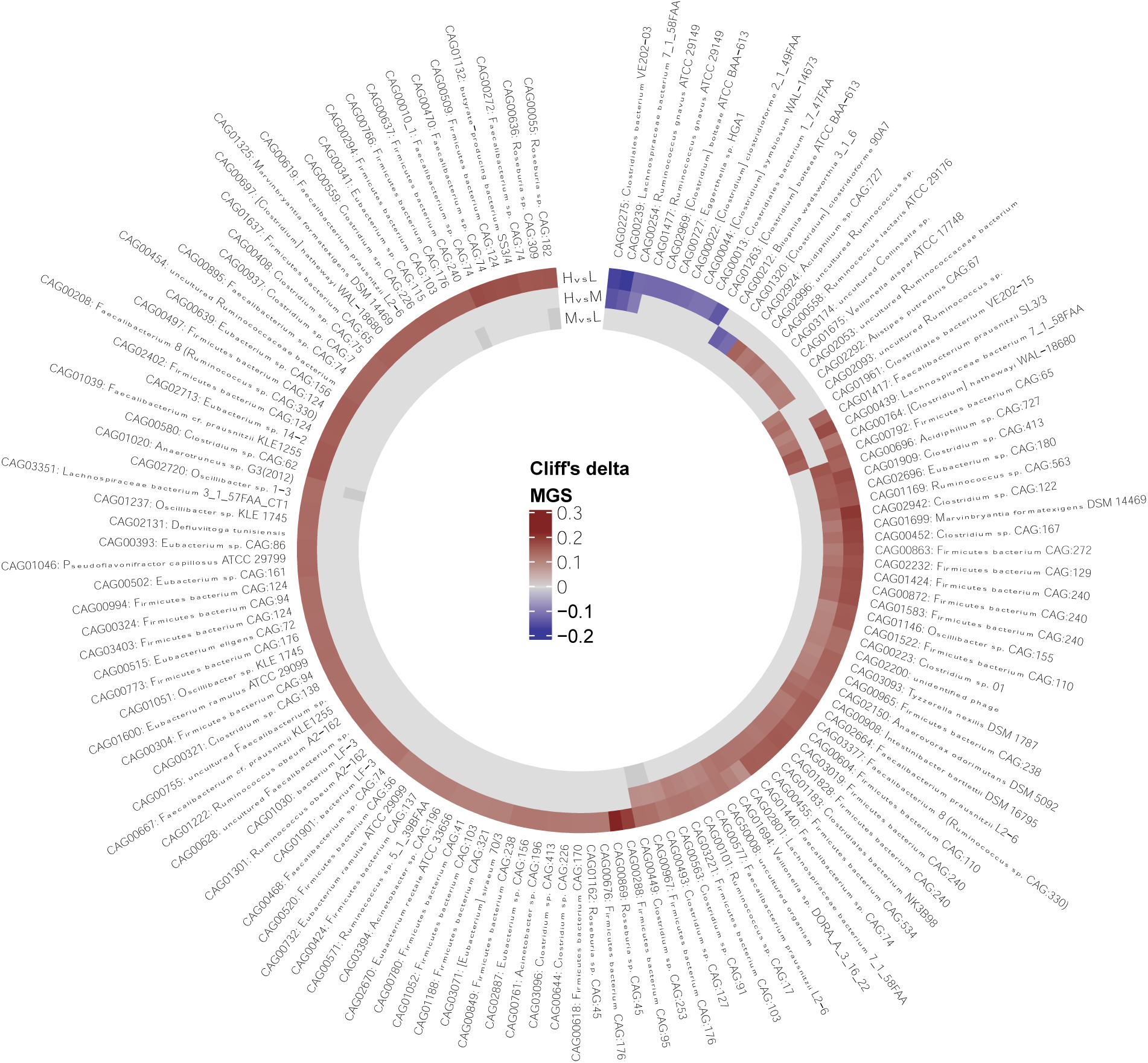
Metagenomic species (MGS) significantly altered between levels of the QASD lifestyle score: Circle plot showing at each layer the effect sizes (cliff’s delta) of 135 MGS with significant differences between levels of the lifestyle scores (HvsL=High vs. Low; HvsM=High vs. Medium; MvsL=Medium vs. Low; Low=QASD 0,1,2,3; Medium = 4; High= 5,6,7) and strictly deconfounded by intake of metformin, statin, PPI intake and Metacardis clinical groups according to metadeconfoundR results (FDR<0.1, OK_sd status and absolute Cliff’s delta effect sizes greater than 0.1). Positive values correspond to higher abundance of the MGS in the reference level of the pairwise comparison (p.ex. in HvsL, Cliff’s delta>0=High abundance in level high of the lifestyle score vs. level Low; Cliff delta<0=Low abundance in level high of the lifestyle score vs. level Low). The full list of 194 MGS with significant differences between levels of the lifestyle scores are provided in Supplemental Table S7.

Regarding metabolomic signatures, elevated QASD score showed a positive association with hippurate, compounds derived from tryptophan metabolism, acyl-cholines, chemical compounds associated to coffee consumption/metabolism or pyrimidine related metabolites. Likewise, reduced levels of the QASD score showed positive association with branched-chain aminoacids and related metabolites, bile-acid metabolites, markers of tobacco consumption and dipeptides (see color-coding in Figure 5, see Supplemental Results and Supplemental Table S9 for extended results about literature evidence around serum metabolomics signatures). Our findings also highlight that the predictive efficacy of these deconfounded serum metabolites surpasses that of the MGS in binary classification of individuals across QASD score levels. The most notable results were obtained with random forest (AUC=70.8) and ternary Predomics models (AUC=69.77) specifically in high vs. low QASD score classification, underscoring the superior performance of serum metabolites (Supplemental Figures 5, 7; detailed results in Supplemental Results).

**Figure 5:**
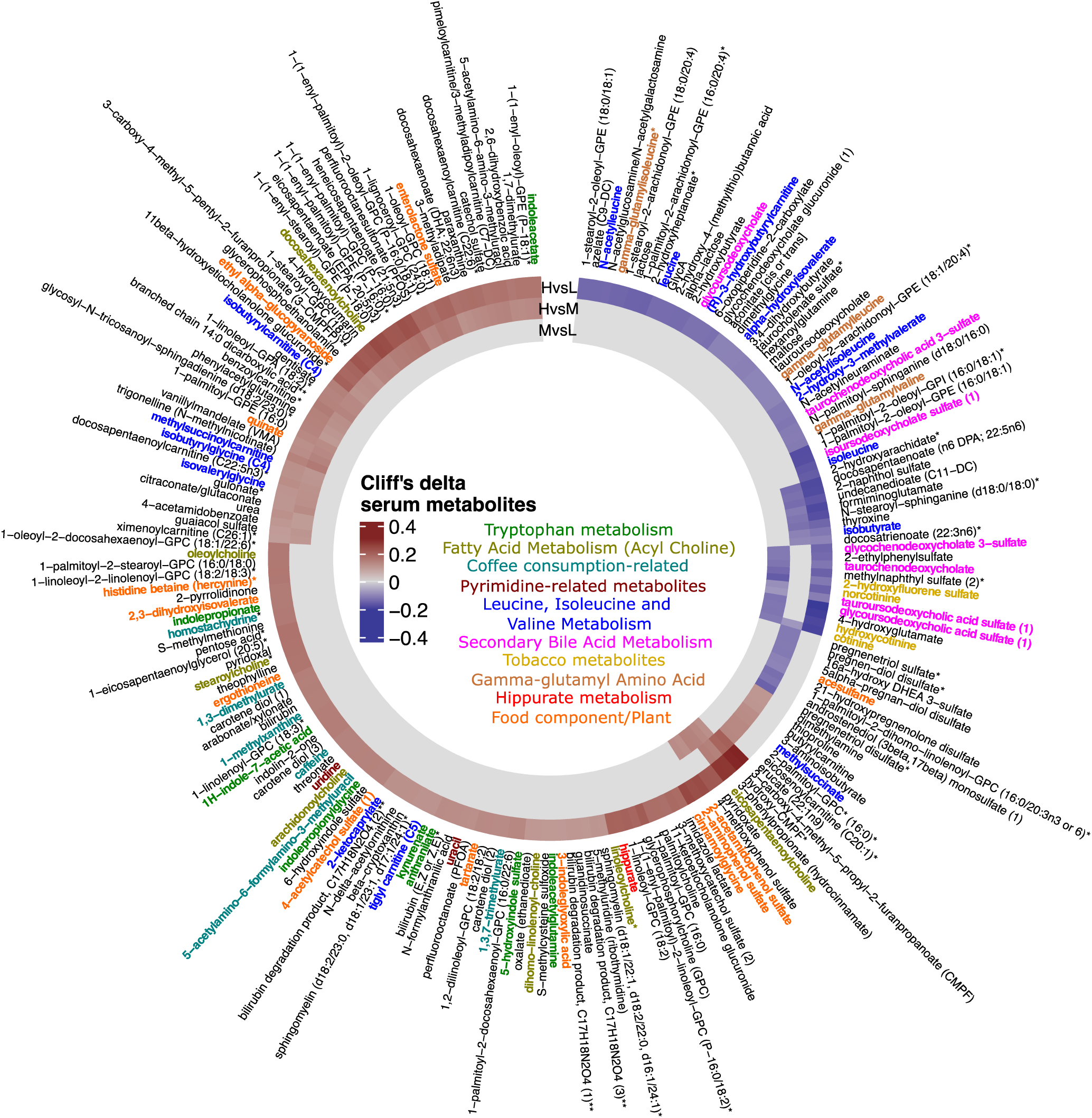
Metabolites significantly altered between levels of the lifestyle score: Circle plot showing at each layer the effect sizes (cliff’s delta) of 204 annotated serum metabolites with significant differences between levels of the lifestyle scores (HvsL=High vs. Low; HvsM=High vs. Medium; MvsL=Medium vs. Low; Low=QASD 0,1,2,3; Medium = 4; High 5,6,7) and strictly deconfounded by intake of metformin, statin, PPI and MetaCardis clinical groups according to metadeconfoundR results (FDR<0.1, OK_sd status and absolute Cliff’s delta effect sizes greater than 0.1). Positive values correspond to higher abundance of the metabolite in the reference level of the pairwise comparison (p.ex. in HvsL, Cliff’s delta>0=High abundance in level high of the lifestyle score vs. level Low; Cliff delta<0=Low abundance in level high of the lifestyle score vs. level Low). The full list of 319 metabolites with significant differences between levels of the lifestyle scores are provided in Supplemental Table S7.

This analysis unveiled MGS and metabolic species distinctly associated with high vs. low levels of the QASD score, emphasizing the superior predictive power of metabolites in delineating these distinctions.

### Gut microbiome strongly mediates interaction between the QASD score and serum metabolomics profile

We conducted the mediation study to investigate whether the metagenome significantly mediates the relationship between the QASD score and individual blood metabolomic profiles. We explored the MGS engaged in this mediation and blood metabolites impacted by this interaction. To do so, we concentrated on selecting features that had demonstrated robust deconfounded associations with the QASD score, including 135 metagenomic species (MGS) and 272 metabolomic signatures with an OK_sd status, FDR<0.1, and an absolute effect size (Cliff’s delta) superior than 0.1.

We examined two primary mediation hypotheses. The first hypothesis investigated the role of MGS as mediators in the relationship between the QASD score and the host’s metabolome, i.e., mediation by the microbiome (Direction 1, Figure 6A). The second hypothesis explored whether the interaction between the QASD score and the host’s metabolome could impact the metagenomic landscape, i.e., reverse mediation (Direction 2, Figure 6A). These hypotheses were tested across all deconfounded pairs of MGS and serum metabolites, considering different levels of the QASD score (high vs. low, high vs. medium, medium vs. low). For inclusion in the analysis, these pairs needed to exhibit a significant association.

**Figure 6:**
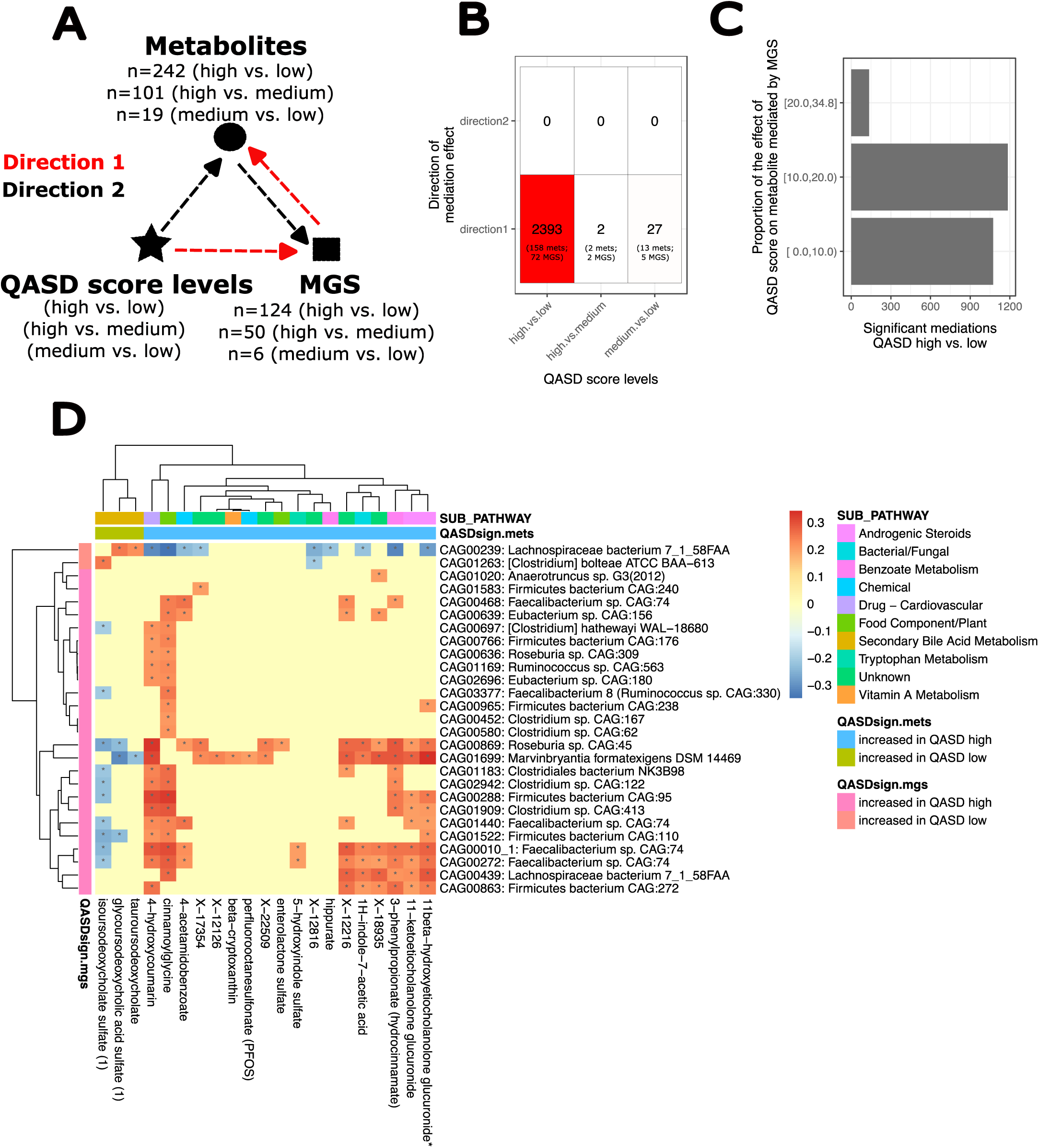
Overview of *in-silico* bidirectional causal mediation analyses between levels of QASD score, metabolites and metagenomic species (MGS) (A) Schematic representation of the two directions tested for causal mediations potentially driven by the QASD score (independent variable). In direction 1, the QASD score would impact serum metabolites (outcome variable) mediated by the abundance of MGS (mediator), whereas in direction 2 the QASD score would impact MGS (outcome variable) mediated by serum metabolites (mediator). Mediation tests were done for all pairwise combinations of MGS and serum metabolites with significant variations between levels of the QASD score retrieved from the drug deconfounding analyses. Mediation analyses were adjusted by center of recruitment, age, gender and intake of metformin, statin and PPI (B) Summary of the number of significant mediations (FDR<0.05 for Average Causal Mediation Effect (ACME), Average Direct Effect (ADE) and Total effect) retrieved in each direction (y-axis) across pairwise levels of the QASD score (x-axis). (C) Decomposition of the 2393 significant mediation relationships in direction 1 (QASD->MGS->Metabolite) observed when considered the extremes of the QASD score (high vs. low) based on the proportion of the total effect of the QASD score on serum metabolite mediated by MGS abundances (y-axis). In 135 of these mediations the mediator (MGS; n=27) drives >20% of the total effect of the QASD score on serum metabolites (n=21) (D) Details of these 135 causal mediations with the highest effect sizes in terms of proportion of the effect driven by the mediator (MGS). The heatmap represents the proportions of the effect of QASD score on serum metabolites (x-axis) that is significantly mediated by MGS (y-axis; *=FDR<0.05 on ACME, ADE, and Total effect). The sign of the effect reflects the sign of the mediator effect (MGS) on serum metabolite levels (positive=MGS and serum metabolite positively associated; negative=MGS and serum metabolite negatively associated). Colors on top and left sides of the heatmap represents the association with QASD score of serum metabolites and MGS respectively as well as main compound sub-pathways.

Our findings strongly supported the first hypothesis. Specifically, for extreme QASD score levels (high vs. low), we found 2,393 significant mediations involving 158 metabolites and 72 MGS, in absence of significant reverse mediation (Figure 6B and Supplemental results). Similar results were noted considering intermediate QASD score levels, but revealing only 2 significant mediations in direction 1 for high vs. medium comparisons and 27 for medium vs. high comparisons of QASD score levels vs. no significant mediation in direction 2 (Figure 6B). These findings emphasize that extremes of the QASD score levels have the most significant impact on an individual’s metagenomic and metabolomic profiles, mainly influencing serum metabolite patterns via potential changes in the microbiome (e.g., MGS abundances). Moreover, mediation effects of MGS abundances on the impact of QASD score on serum metabolites ranged from 2.5% to 34.8% of the total effect. Particularly noteworthy were the 135 high-impact mediations, where MGS abundances explained over 20% of the QASD score’s effect on serum metabolites (Figure 6C). This set of mediations involved 27 MGS and 21 serum metabolites, shown in Figure 6D. These mediations highlighted potential mechanisms through which the gut microbiome may influence chemical compound levels, encompassing both positive and negative relationships according to the sign of the mediator effect on serum metabolite levels (Supplemental Figure 8, Supplemental Results).

One of the most pronounced positive mediation effects was evident with 11 beta-hydroxyetiocholanolone glucuronide, a glucuronated steroid conjugate. In individuals with a high QASD score, 34.3% of this steroid increase was positively mediated by the elevated abundance of CAG01699: *Marvibryantia formatexigens* DSM 14469 (Figure 6D). This lineage is known for hosting genes encoding different types of beta-glucuronidases, which hydrolyze dietary beta-glucuronides^42,43^, suggesting a potential mechanism for this positive mediation. *Faecalibacterium* lineages, which are also more abundant in subjects with a high QASD score, may also play a role in these mediating effects. These lineages contribute to increase the levels of glycine-conjugated compounds, such as cinnamoylglycine or 3-phenylpropionate (Figure 6D), known as serum biomarkers associated with high microbiome diversity^44,45^. The microbial origin of these compounds has been demonstrated in previous mouse experiments, as evidenced by cinnamoylglycine ^46^. The positive mediation observed between dietary beta-cryptoxanthin, a carotenoid primarily found in fruits and vegetables, and CAG01699: *Marvinbryantia formatexigens* DSM 14469 (Figure 6D) can be interpreted as the positive influence of nutritional compounds on gut microbiome species, as it has also demonstrated dose-dependent probiotic effects^47^.

In individuals with low QASD scores, positive mediations involved secondary bile acids. For instance, isoursodeoxycholate sulfate had 25.2% of its increase in individuals with low QASD scores directly mediated by the increased abundances of CAG01263: *[Clostridium] bolteae ATCC BAA-613* (Figure 6D). To support a potential mechanistic explanation for this mediation, the genome-scale metabolic models of *C. bolteae* from the AGORA2 repository, including the ATCC_BAA_613 strain, contain bile salt hydrolases capable of deconjugate the primary bile acids taurochenodeoxycholate and glycochenodeoxycholate by releasing taurine and glycine moieties respectively, from which the isoursocholate precursor ursodeoxycholate are produced ^48,49^. As an instance of negative mediation, the rise in isoursocholate sulfate among individuals with low QASD scores was also mediated by reduced abundance of *Faecalibacterium* lineages (Figure 6D).

Taken together, these results reveal intricate ecological interplays in the gut microbial ecosystem, which may be crucial in determining the serum metabolome signatures linked to the novel QASD lifestyle score among individuals with cardiometabolic disorders.

## Discussion

Our study sheds light on the intricate and complementary impacts of lifestyle on the gut microbiome and host metabolic phenotypes, including blood metabolome, underlining the multifaceted nature of these interactions^50^. In this extensive investigation of European populations, encompassing both healthy individuals and individuals with a broad spectrum of cardiometabolic diseases, we utilized rigorous analytical methods to unveil the significant relationships between a novel composite lifestyle score (QASD) and fecal microbiome richness and composition and host phenotypes including molecular phenotype (e.g., metabolome). This analysis highlights the substantial role played by specific gut microbiome species in mediating relationships between this composite score and a number of metabolites, as well as the mediating role of the gut microbiome status (GMGR) in the impact of QASD score on glucose metabolism phenotypes.

Our study shows that the gene richness and more largely the composition of the gut microbiome, including the enterotypes, are not only associated with a variety of food items, nutrients, polyphenols and vitamins but also to broader dietary patterns, physical activity levels, and smoking status, that can be combined. As highlighted by the American Heart Association^16^, comprehensive approaches with composite scores is critical in nutrition research since it provide an integrated view of lifestyle with individual patterns. We, here, not only use combined dietary score but also propose to combine with important features of cardiometabolic risks as physical activity and smoking. We reveal intricate relationship between integrated lifestyle patterns in QASD score, the microbiome and metabolome remains generally underexplored in population with cardiometabolic disorders ^7,51,52^.

We notably observed a consistent association between a high QASD score levels and the microbiome compositional space corresponding to the *Ruminococcus* enterotype whereas a low QASD and a high dietary inflammatory index (aDII) were linked to the Bact2 enterotype. These associations of the QASD score and the dietary inflammatory index with the gut microbiome enterotypes were replicated in an independent cohort from the GutInside study with overweight and obesity. Whereas enterotype definition has been controversial from their first definition based on beta-diversity ^53–55^, there is a consensus that enterotype exists in the context of a continuous landscape of microbiome compositional variation ^56^. Studies also propose a more complex scenario of 24 enterotypes using the DMM approach on pooled metagenomic datasets from 86 studies ^57^ or to 5 community types or enterosignatures using non-negative matrix factorization approach ^58^. Despite these variations, the presence of a Bacteroides-enriched composition characterized by low microbial diversity is common and consistent associations were found between the Bact2 enterotype and a higher BMI, elevated inflammatory blood markers and a lack of statin consumption in multiple cohorts including both the MetaCardis study and our confirmatory population (e.g. GutInisde.^2,30^).

Crucially, our results reveal that the associations between dietary diversity and gut microbial gene richness are independent of food quality. Our work adds to previous observations made in relatively healthy large-scale populations like the PREDICT and Flemish Gut^6,7^ by highlighting the impact of dietary diversity on the microbiome composition which extends to dietary components, generally considered unhealthy in nature. The positive association of physical activity variables with microbial gene richness is also in line with observations made in relatively healthy populations like the US Arrivale cohort^36^.

Our proposed QASD score, that combines the variables most strongly associated with microbial richness (e.g. including alcohol as part of the aHei) demonstrates a more substantial effect size of its impact of microbiome composition across the entire population and within MetaCardis pathology subgroups, relative to its singular components. Indeed, the influence of individual dietary elements or distinct lifestyle factors in accounting for changes in the gut microbiome is usually minimal, a trend similarly observed in other populations^51,59,60^.The four key variables (e.g., dietary quality, dietary diversity, physical activity levels, and smoking status) of the QASD score may exert partially non-redundant effects ^49^. Noteworthy, when we examined the most influential lifestyle variables which had non-redundant associations with microbiome composition, we found that they collectively accounted for less than 2% of the microbiome’s variability. This fraction increase when other known clinical covariates are considered, but importantly we observed that the main components of the QASD score (aHEI and smoking status) significantly contributes to explains a non-redundant part of the microbiome compositional variation that is not explained by these clinical covariates. This underscores the multifaceted nature of the microbiome, influenced by a myriad of lifestyle, food-related factors, genetics (not explored here) or and individual host phenotypes^6,61,62^.

Nonetheless, as we delved deeper into the metagenomic and metabolomic signatures associated with our QASD score, we unearthed compelling associations. QASD score was strongly associated to 135 decounfounded metagenomic species and 272 metabolites, including metagenomic species enriched in individuals with high QASD score that were previously suggested to be part of healthy microbiome signatures like *F. prausnitzii, E. eligens, Oscillibacter*, or *Roseburia* genera influenced by nutritional changes or depleted in chronic diseases^7^. Conversely, lower QASD scores were associated with metagenomic species previously linked to inflammation or potentially considered markers of poor health. These included lineages like *C. bolteae* and *R. gnavus*, which displayed correlations with circulating markers of increased fasting and post-prandial inflammation in the PredictUK study^7^.

Importantly, the interaction between microbiome and metabolic health can be bidirectional, making it essential for future research to explore in depth these relationships and find relevant mediators. As such mediation analyses has been previously applied to evaluate the interplay between dietary factors, plasma metabolomics and gut microbiome features^61^, to show for example a mediation of microbial vitamin production on the influence of fruit intake on diabetes risk^63^, or to evaluate whether some metabolites can mediate the microbial impact on host phenotypes^64^. Here, using the mediation analysis, we demonstrated that a major fraction of MGS, played a significant mediating role in the impact of our lifestyle score, on individual metabolite profiles (ranging from 2.5% to nearly 35% of the effect on serum metabolomics profiles). Among these, high-impact mediations explaining more than 20% of the QASD score’s effect on serum metabolome involved 27 MGS and 21 metabolites. MGS include some the above cited MGS like the increase in *C. bolteae* in individuals with low QASD score, which strongly mediates the increases in secondary bile acids like isoursodeoxycholate sulfate, a serum biomarker associated to post-prandial lipemia, inflammation and worse hepatic function^65^.

Previous mediation analyses on individual dietary variables derived from FFQ data, the microbiome and plasma metabolomics have evidenced complex mediation relationships where a dietary factors impact plasma metabolome mediated by microbiome features (as we observe with our QASD score) like a mediating role of *Ruminococcus species vSV* on the effect of fruit consumption on plasma levels of urolithin B, but also multiple mediations in the opposite direction, where a dietary factor impacts microbiome features also mediated by plasma metabolites, like a mediating role of plasma hippurate on the impact of coffee intake on the archaeon *Methanobrevibacter smithii*^61^.

As such when considering mediation analysis with GMGR, we found that GMGR mediated the relationships between glucose homeostasis biomarkers and the QASD score, but with a different picture than metabolome exploration. Indeed, clinical variables also mediated the association between lifestyle and GMGR, highlighting the bidirectional relationships. As such, fasting glucose and insulin sensitivity markers, rather than BMI, appeared to be particularly associated with lifestyle and the gut microbiome. This is in accordance with previous research^66^. Interventional studies on personalized diets have shown that gut microbial pathways and species, including different *F.prausnitzii* subgroups, mediates the effects of components of the diets on metabolic health ^67,68^. Other studies in the LifeLinesDEEP cohort have shown a mediatory role of microbial B1/B2 vitamin production on the influence of fruit intake on diabetes risk^63,64^. This observation is in agreement with recent findings showing that the gut microbiome is a modulator of the beneficial association between a Mediterranean diet and cardiometabolic disease risk, using prospective data in 307 male participants^69^, as well as with studies that have shown a significant mediation of microbiome diversity and composition on the effect of green Mediterranean diet in the reduction of cardiometabolic risk^70^. Recent studies have shown that the impact of the microbiome diversity on metabolic health is also mediated by the diversity of serum metabolites itself^44^, emphasizing further the interplay of the microbiome and metabolome in human metabolic health.

As limitations of our study, to perform these explorations, we used mostly long-term food habit (e.g., FFQ) and future research should explore using other standards for dietary assessment, such as dietary 24-hour recalls. Moreover, individual optimization of the composite score should be undertaken in the future to find individual lifestyle variables to improve gut microbiome-metabolic health interaction. Also, the study paves the way to mechanistic exploration to examine the relevance of causal mediation relationships inferred from statistical analyses, notably for MGS or metabolite candidates involved in the most significant mediations.

Overall, our findings shed new light on the importance of gut microbiome in mediating the intricate and complex relationships between individual lifestyle pattern, and the host metabolic phenotypes in people with cardiometabolic disorders. Comprehensive understanding of these interplays could help designing recommendations, especially for at-risk populations, by combining various nutrition and lifestyle factors with significant impact on gut health, enhancing cardiometabolic health strategies.

## Data availability

Raw shotgun sequencing data that support the findings of this study have been deposited in the European Nucleotide Archive with accession codes PRJEB37249, PRJEB38742, PRJEB41311, PRJEB46098 and PRJEB71898, with public access. Metabolome data have been uploaded to Metabolights and MassIVE with respective accession numbers—that is, serum NMR and urine NMR with accession number MTBLS3429.

## Acknowledgments

We would like thank the subjects for their participation in the MetaCardis study and particularly patient associations (Alliance du Coeur and CNAO) for their input and interface, the nurses, technicians, clinical research assistants and data managers from the Clinical investigation platform at the Institute of Cardiometabolism and Nutrition for patient investigations and the Clinical Investigation Center (CIC) from Pitié-Salpêtrière Hospital for investigation of healthy controls. Quanta Medical provided regulatory oversight of the clinical study and contributed to the processing and management of electronic data.

## Author contributions

SA, EB, MCD and KC conceived and designed the project. SA and EB assisted with data management, curation, performed dietary and metagenomic analysis and SA, EB and KC wrote the manuscript. SKF assisted with data management, curation, deployment of confounder-adjusted analysis tools and results interpretation, and further provided feedback on the manuscript. TLR, MCD, JDZ, KC assisted with results interpretation, and further provided feedback on the manuscript. TS and GAll authors commented and edited the manuscript. KC is the guarantor who accepts full responsibility for the work and/or the conduct of the study, had access to all data, and controlled the decision to publish.

## Funding

This work was supported by European Union’s Seventh Framework Programme for research, technological development and demonstration under grant agreement HEALTH-F4-2012-305312 (METACARDIS). Funding supports were also obtained from Leducq Foundation(17CVD01), JPI-Microdiet study (2017-01996_3). Part of the work was supported by a grant from the Deutsche Forschungsgemeinschaft (DFG): SFB 1052 (project B1), the Fondation pour la Recherche Médicale (FDT201904008276, FDT202106012793), and the French Agency of Research (ANR-NUTRIM_CHECK, ANR-DeepIntegromics). SKF is supported by the Deutsche Forschungsgemeinschaft (DFG, German Research Foundation; SFB1365 and SFB1470) and the Deutsches Zentrum für Herz-Kreislauf-Forschung (DZHK).

## Conflict of Interest

No conflict of interest was reported in relation with this study.

## Methods

### Study cohort and sample acquisition

This study is based on the European MetaCardis cohort (http://www.metacardis.net/), initially designed to study the relationships between the different stages of the disease and the gut microbiome. The cross-sectional European MetaCardis study cohort covered a wide range of metabolic and cardiac phenotypes of 2214 participants that were recruited between 2013 and 2015 in Denmark, Germany and France. The study cohort, including protocol information, exclusion criteria, definition of the groups, biochemical analyses, collection of anthropometric and clinical data, were extensively described previously ^2,3,29,71^.

Briefly, for the purpose of the present study we focused on a subset of 1,643 individuals, including healthy normal-weight individuals (BMI <25 kg/m²) as well as individuals with non-exclusive metabolic conditions such as type 2 diabetes, metabolic syndrome, obesity and cardiac disease, as summarized on the flow chart of Supplemental Figure 9 and described in the supplementary Table S1. The study protocol was approved by the Ethics Committee at the Medical Faculty at the University of Leipzig, Germany (application number: 047-13-28012013), the ethical committees of the Capital Region of Denmark (H-3-2013-145) and the ethics committee ‘Comité de Protection des Personnes’ (CPP) Ile-de-France III no.IDRCB2013-A00189-36 in France and was registered at https://clinicaltrials.gov/ (NCT02059538). The observational cohort design complied with all relevant ethical regulations, aligning with the Helsinki Declaration and in accordance with European privacy legislation. All participants provided written informed consent.

As a cohort of replication for people with overweight and obesity, we used data from the GutInside study as described in Table S2 ^30^. Subject were recruited in France from September 2018 to January 2020 in different regions and the population was composed of 433 individuals with overweight or obesity (BMI ≥ 25 kg/m²). While there were involved in a dietary intervention program, we here only examined baseline data for the purpose of this work. Similar questionnaires were used to evaluate lifestyle aspects.

### Clinical, anthropometric variables and definition of study groups

Groups were defined along international definitions of disease, with overweight and obesity assessed according to the WHO criteria (BMI ≥ 25 and BMI ≥ 30 kg/m², respectively). The metabolic syndrome (MS) was defined based on the International Diabetes Federation 2005 Consensus Worldwide Definition of the Metabolic Syndrome (http://www.idf.org/metabolic-syndrome), e.g. waist circumference> 94 cm in men and> 80 cm in women, and any two of the following four factors: i) triglycerides levels ≥ 1.7 mmol/L or treatment for lipid abnormality (statin and/or fibrate or ezetimibe); ii) high density lipoprotein (HDL) cholesterol< 1.03 mmol/L for European men and 1.29 mmol/L in European women and/or treatment for lipid abnormality; iii) abnormal blood pressure (BP): systolic BP ≥ 130 mmHg and/or diastolic BP ≥ 85 mmHg or patients taking anti-hypertensive treatment and, iv) abnormal fasting plasma glucose ≥ 5.6 mmol/L or prevalent T2D. T2D status was defined using the American Diabetes Association (ADA) definition: fasting glycemia> 6.9 mmol/l and/or 2h values during an oral glucose tolerance test> 11 mmol/l and/or hemoglobin A1c (HbA1c, glycated hemoglobin) ≥ 6.5% and/or use of any anti-diabetic treatment (American Diabetes Association 2018). Hypertension status was defined according to the American College of Cardiology and American Heart Association^72^. For obesity specifically, participants were recruited into two groups: group 2A, consisting of individuals with mostly grade II obesity (BMI≥ 35) none of whom had T2D or previous cardiovascular conditions, whereas group 2B consisted mostly of individuals with grade III obesity (BMI ≥40), who were eligible for bariatric surgery. T2D was not an exclusion criterion for this particular group (in contrast to group 2A) and patients from group 2B had overall more severe metabolic impairments than those in group 2A. Individuals with heart failure were defined according to the American College of Cardiology, American Heart Association and the Heart Failure Society of America ^73^. We combined the group 2A and 2B in this study for descriptive purposes. Our group of participants in the present study with coronary heart disease included patients with first events of acute coronary heart diseases, chronic coronary artery disease with heart failure (defined as a left ventricular ejection fraction ≥45%) or without heart failure (<45%). We excluded participants suffering from heart failure related to an origin other than coronary artery disease.

Weight, height and waist circumference were assessed during the clinical inclusion visit according to standardized procedures using the same scales and units. Body fat mass and fat-free mass were measured via bioelectrical impedance analysis. Systolic and diastolic blood pressure were measured using a mercury sphygmomanometer (measures were taken three times on each arm, and the mean of the last 2 measurements in the right arm was used for analyses). Moreover, a detailed list of prescribed medications as well as the patient’s medical history were gathered. We have carefully cleaned and integrated each drug treatment according to its class of molecules, as described in this study ^29^.

### Dietary intake and lifestyle data assessment

#### General questionnaire and database

Dietary data for the MetaCardis cohort was collected via a web-based validated food-frequency questionnaire that was adapted to the cultural habits of each of the countries where recruitment took place. FFQs display a preselected list of foods for which individuals are asked to indicate the frequency of food consumption over a period of time and sometimes to state the average amount consumed. The MetaCardis FFQ was based on the validated European Prospective Investigation of Cancer (EPIC)-Norfolk FFQ and the content was based on several relevant European FFQs. A validation study against repeated 24h24h- dietary records among 324 French MetaCardis participants has indicated a good validity for micronutrients^31^. Portion size and nutrient composition were derived from national food consumption surveys and food composition databases.

Since the initial version, several compounds and nutrients have been added to the initial composition table, allowing us to more accurately study compounds such as sub-classes and total of polyphenols, derived from the updated Phenol-explorer table^74^ or nutrients/metabolites related to the updated USDA database (https://fdc.nal.usda.gov/), FooDB (https://foodb.ca/) and the Danish national food database (DTU food database: https://frida.fooddata.dk/) for biotin intakes ^3^. We have carefully derived and integrated each nutrient or metabolite of interest in a consortium internal composition table, thanks to dietary matching and detailed recipe decomposition work carried out by dieticians and experts in the field.

For each subject, basal metabolic rate (BMR) was estimated using Harris and Benedict equation. Subjects who overtly under- or over-reported energy intake defined as <0.5*BMR or >3.5*BMR were excluded from dietary analyses (<10% of the subjects with available nutritional data).

Data on physical activity (PA) were collected using the Recent Physical Activity Questionnaire (RPAQ). This questionnaire assesses the habitual physical activity in the past month takes into account 4 physical activity domains (home, work, travel, and leisure time) and has been validated against doubly labelled water ^32^ . Physical activity data for each participant are computed both as hours per week as metabolic equivalent of task (MET) minutes per week, using a published compendium of physical activities and their associated MET values ^75^. One MET corresponds to energy expended when sitting at rest and is set at 3.5 ml/min/kg oxygen consumption ^75^.

The data on smoking status were obtained through several questions on the smoking or non-smoking status of the subject, on the number of cigarettes smoked, the date of cessation or resumption of smoking, passive smoking. We have used the categorical variable on smoking status presented in Supplemental Table S1 (non-smoker/ancient smoker/passive smoker/current smoker) recoded in 2 categories (non-smoker/smoker) for QASD score calculations (see below). The initial variable “smoking status” coded in categorical (non-smoker/ancient smoker/passive smoker/current smoker) was cross checked with the quantitative data of the number of cigarettes per day, the date of start or stop of smoking and other data regarding passive smoking, thereby confirming the smoking status (non-smoker/smoker).

### Foods groups and nutrient content

The initial responses to the FFQ are frequency choices that indicate consumption units per month, week or day for each 160 food items. These frequency choices are then converted to daily frequencies and finally to food group intakes in g/d and to nutrient intakes in g/d, mg/d or mcg/d – based on information on portion size (i.e., grams contained in one portion) and nutrient content (i.e., nutrients contained in one portion). This information was taken from nationally relevant food composition databases.

Consumption of food groups was obtained by grouping the 160 food items into nutritional interest groups. The 22 initial groups of the MetaCardis study were broken down into 42 subgroups (shown in Supplementary Table S4). Dietary diversity scores were based on these 42 subgroups, according to *ad hoc* criteria. In sensitivity analysis, we constructed these same dietary diversity scores by removing “liquid” foods from the diet, i.e., hot drinks, fruit juices, sugary drinks, alcoholic drinks and milk because the quantities consumed can unbalance the diversity scores based on the uniformity of consumption (Simpson index).

### Dietary “inflammatory potential” of the diet

The “inflammatory potential” of the diet was determined using the alternative Dietary Inflammatory Index (aDII) ^76,77^, an alternative version of the original Dietary Inflammatory Index (DII) ^78^, developed by Cavicchia et al. ^79^ and following the methodology initially provided by van Woudenbergh et al. ^80^. Briefly, the ADII included 34 food items, which were standardized after adjustment for energy intake using the residual method developed by Willett et al. ^81^ to reduce the variation in dietary intake resulting from differences in physical activity, body size, and metabolic efficiency ^82^. Each item was then multiplied by a proposed coefficient ^78^, and items were summed to create the aDII score. Higher aDII values reflect a more proinflammatory diet.

### Dietary quality scores

The Alternative Healthy Eating Index (AHEI) and Dietary Approaches to Stop Hypertension (DASH) scores were computed to evaluate overall dietary quality, two *a priori* scores, taking into account the existing correlations between the different components of the diet developed in order to overcome the limitations linked to approaches focusing only on food or nutrient groups ^83^ and based on foods and nutrients predictive of chronic disease risk ^84,85^. As a measure of healthy US-style eating, the AHEI-2010 assigns 0–10 points to each of the 11 dietary components based on the portion size ^83,84^. The DASH score contains 8 components, each of which receives 1–5 points according to its consumption quintile ^86^.

### Dietary diversity and variety scores

Several dietary diversity and variety groups were constructed, depending on the groups retained in the method of construction (42 groups aggregated, with or without “liquid” foods, presented in Supplemental Table S4) and the method retained for the estimation (count-based or evenness-based estimation).

First, we used count measures of the diversity, whereby the number of consumed food items and food groups is recorded. Count measures focus on counting different food groups and subgroups; the distribution of consumed food quantities is not taken into account. Nevertheless, research studies have been limited by count-based measures and inconsistencies in the number and types of foods groups considered in these estimations ^16^. To overcome these limitations, we have systematically varied the number of foods groups in sensitivity analyses, in particular removing liquid foods from the diet. Secondly, we have used measures of the diversity in terms of number as well as distribution of different food items, using the Berry-Index, which is also known as the Simpson-Index^87^, indexes also used to assess metagenomic or ecological diversity for each measure of the diversity considered in the count-based at the first step. This score increases if the food items are more equally consumed rather than being more concentrated.

We calculated the HFD score designed and published by Drescher et al. ^40^. This index could be interpreted as a healthy food diversity indicator, i.e. the diversity of the healthy products quality (dietary diversity of “considered” health products) ^40^. Finally, we also calculated the normalized composite dietary and variety score of the diet ^38,39^, a score that combined diversity and variety (simple counts, normalized on a scale of 0 to 100).

All these scores account for different aspects of dietary diversity and variety. For example, some scores take into account the entire diet (identified by the word “diversity all diet” in Figure 1B), while others focus on specific groups of the diet, e.g., food items (identified by the word “variety” in Figure 1B). We used diversity and variety scores based on simple counts in consumption groups (identified by the word “count” in Figure 1C) and scores taking into account the evenness of consumption within groups (identified by the word “Simpson” in Figure 1C). Simple counts-based scores were calculated by assigning 0 points for each food group (diversity scores) or food considered in the group (variety scores) if the person did not consume the food or group in question and 1 point if she consumed it (consumption >0 grams). Thus, total diet diversity was a score ranging from 0 to 42 initially groups derived from the FFQ (see the initial 42 groups in Supplemental Table S4).

Simpson based scores were calculated using food groups (diversity) or foods in a given food group (variety) as different species, by analogy with (microbial) species used to calculate ecological indexes to measure the diversity of a given environment. Simpson indexes take into account the quantity of species present but also their abundance (equity).

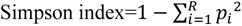

R: richness: number of species considered, p*i* =relative abundance of the species (share of product *i* in the total amount of food consumed).

By subtracting this score from 1, we therefore obtain an index bounded between 0 and 1, 0 corresponding to the minimum diversity and 1 to the maximum (in equal shares of all products considered).

To overcome inconsistencies in the number and types of foods or food groups considered we have performed sensitivity analyses of these scores, varying the number of groups used in taking these calculations into account. In particular, when assessing total diet diversity, we created scores that did not take liquid food into account (See supplementary Table S4 for additional food groups excluding beverages and milk variables).

### The QASD (Quality Activity Smoking Diversity) lifestyle score construction

A new lifestyle score was designed considering the quality of the diet, total physical activity, smoking status and dietary diversity. The score was calculated based on the tertile values of variables corresponding to: food quality (aHEI: 0 point for low tertile, 1 point for medium tertile and 2 points for high tertile), dietary diversity of the all diet, without beverages (Simpson Index) (0 point for low tertile, 1 point for medium tertile and 2 points for high tertile) and total physical activity in MET-h.week^-1^ (0 point for low tertile, 1 point for medium tertile and 2 points for high tertile). For smoking status 0 point was given for smokers and 1 point for non-smokers. Taking together these variables gives a theoretical score between 0 and 7. We then grouped the modalities 0, 1 and 2 and 6 and 7 to correct for low numbers in some categories during the stratifications, giving from the lowest to the highest lifestyle score, 5 modalities: “0-1-2”, “3”, “4”, “5”, “6-7” for Figure 1,3 and Supplemental Figures 1-3 and 3 modalities “0-1-2-3”, “4”, “5-6-7” for Figures 2,4,5 and Supplemental Figures 4-8.

### Extraction of fecal genomic DNA and gut microbiota sequencing

Gut microbiota sequencing in the MetaCardis cohort was performed as previously described^2,29,71^. Briefly, participants collected fecal samples within 24 hours before enrollment. Samples were either stored immediately at −80°C or briefly conserved in home freezers, before transport to the laboratory where they were immediately frozen at −80°C following guidelines53. Total faecal DNA was extracted following the International Human Microbiome Standards (IHMS) guidelines (SOP 07 V2 H) and sequenced using ion-proton technology (ThermoFisher Scientific) resulting in 23.3 ± 4.0 million (mean ± SD) 150-bp single-end reads per sample on average. Reads were cleaned using Alien Trimmer (v0.2.4)^88^ in order to remove resilient sequencing adapters and to trim low quality nucleotides at the 3’ side (quality and length cut-off of 20 and 45 bp, respectively). Cleaned reads were subsequently filtered from human and potential food contaminant DNA (using human genome RCh37-p10, Bos taurus and Arabidopsis thaliana with an identity score threshold of 97%). Filtered high-quality reads were mapped with an identity threshold of 95% to the 9.9-million-gene catalogue using Bowtie2 (v.2.3.4)^89^ included in the METEOR v3.2 software^90^ (https://forgemia.inra.fr/metagenopolis/meteor). A gene abundance table was generated by means of a two-step procedure using METEOR. First, the uniquely mapping reads (reads mapping to a single gene in the catalogue) were attributed to their corresponding genes. Second, shared reads (reads that mapped with the same alignment score to multiple genes) were attributed according to the ratio of their unique mapping counts. The gene abundance table was processed for rarefaction and normalization and further analysis using the R package MetaOMineR^91^. To decrease technical bias due to different sequencing depth and avoid any artefacts of sample size on low-abundance genes, read counts were rarefied. The gene abundance table was rarefied to 10 million reads per sample by random sampling of 10 million mapped reads without replacement. The resulting rarefied gene abundance table was normalized according to the FPKM (fragments per kilobase of transcript per million mapped reads) strategy (normalization by the gene size and the number of total mapped reads reported in frequency) to give the gene abundance profile table and binned by functional and phylogenetic categories as carried out within the MOCAT2 framework^92^. A total of 3463 metagenomic species (MGS; co-abundant gene groups with more than 50 genes corresponding to microbial species) were clustered from 1,267 human gut metagenomes used to construct the 9.9- million-gene catalogue (reference), as described previously^93^. MGS abundances were estimated as the mean abundance of the 50 genes defining a robust centroid of the cluster (if more than 10% these genes gave positive signals). Microbial gene richness (gene count) was calculated by counting the number of genes that were detected at least once in a given sample, using the average number of genes counted in ten independent rarefaction experiments.

### Quantitative metagenomic profiling of GutInside cohort

Gut microbiome sequencing of fecal samples from 433 individuals of the GutInside study (reference Biomedicines) was performed by GeneWiz (https://www.genewiz.com/) using Illumina HiSeq platform, resulting in 23.17±5.1 million read pairs (mean ± std. deviation) per sample. Raw reads were processed with NGLess^94^ for quality trimming (minimum read quality=25; minimum read length=40), host contaminant removal vs. reference human genome (min. identity=90%, min. match size=45), alignment filtered reads over the 9.9-million gene catalogue (reference) (min. identity=95%, min. match size=45), and generation of gene abundance table with the *dist1* metric of NGLess, equivalent to the counting strategy described above for the Metacardis cohort (first the genome abundances are computed from unique mapped reads and then are corrected by the multiple mapped reads weighted by the coverage of unique mapped reads). The gene abundance table was processed for rarefaction and RPKM normalization with MetaOMineR^91^ R package. The raw gene abundance table was rarefied to 10 million reads per sample and the rarefied gene abundance table was normalized according to the FPKM strategy, from which MGS abundances were computed as described above in Metacardis cohort. From NGLess filtered reads, genus-level abundance profiles in the mOTU space were generated with mOTU v2.6.1^95^, from which enterotype classifications was performed after rarifying to 4000 counts per sample using the Dirichlet Multinomial Mixture (DMM) method^55^ and implemented in the Dirichlet Multinomial R package (Dirichlet-Multinomial Mixture Model Machine Learning for Microbiome Data. R package version 1.28.0). The minimum Laplace metric was used as criteria to select the best stratification of the cohort^55^, which was at 4 discrete microbiome compositions that corresponded to the Rum, Prev, Bact1 and Bact2 enterotypes described in the Metacardis cohort^41^.

### Analyses of microbial diversity and community structure

The estimation of the explanatory power of nutritional features regarding relative microbiome profiles derived from MGS abundance data was performed using univariate and multi-variate stepwise distance-based redundancy analyses (dbRDA) as implemented in the R package *vegan* (Community Ecology Package. R package v.2.2–1)^96^. Redundance filtering of collinear lifestyle variables were done previously to these analyses with the R package FMradio^97^; threshold=0.9 on Spearman pairwise correlation matrix). Microbiome inter-individual variation was visualized by principal coordinates analysis using Bray–Curtis dissimilarity on the relative abundance matrix derived from MGS abundance data. Environmental fitting of nutritional covariates with significant impact on microbiome composition based on dbRDA analyses over PCoA ordination from Bray-Curtis inter-sample dissimilarity matrix was computed with *envfit* function of *vegan* R package. Associations of nutritional variables with enterotype status were tested with logistic regression analyses adjusted by age, gender, center, BMI, and the intake of metformin, statin and PPI.

### Metabolomic profiling

We used untargeted UPLC–MS data generated by Metabolon, as described in previous study^71^. Briefly, serum samples were extracted and profiled by Metabolon using a UPLC–MS-based methodology98. Annotated metabolites and unknown features (denoted X-00000) were identified by comparing sample features with ion features in a reference database of pure chemical standards and previously detected unknowns, followed by detailed visual inspection and quality control, as reported99. For all metabolomic assays, we randomized the sample preparation order across the whole study so that each sample preparation batch included samples from all study groups. For MS untargeted assays, median batch correction was performed by adjusting batch-wise study sample variable medians according to a scalar derived from adjusting pooled reference sample medians, so that pooled reference sample medians are identical across all batches. The randomized sample preparation batches were also tested for association with study groups using univariate statistics (Fisher’s exact test or Kruskal–Wallis test), and P > 0.05 was observed across all methods (GC–MS, Fisher’s exact test, P = 0.23; UPLC–MS targeted, Fisher’s exact test, P = 0.12; and UPLC–MS untargeted (Metabolon), Fisher’s exact test, P = 0.65). In addition, NMR run order exhibited a Kruskal– Wallis P = 0.49.

### Drug deconfounding analysis

The pipeline was used to assess to what extent observed differences among pairwise levels of the QASD score decomposed into 3 levels (high vs. medium, high vs. low, medium vs. low; Low=QASD 1,2,3; Medium = 4; High=5,6,7) between study participants in microbiome, metabolome and bioclinical feature abundance are confounded, in the sense of being consequences of other (treatment or risk factor) variables different among the groups more so than characteristic of the specific phenotype itself. We employed the post hoc filtering approach implemented in the R package metadeconfoundR (version 0.1.8; see https://github.com/TillBirkner/metadeconfoundR or https://doi.org/10.5281/zenodo.4721078) that was devised within the MetaCardis consortium^29,71^. The pipeline has two steps. In the first step, all associations between -omics features and the set of independent variables (disease status, drug treatment status and risk markers, including age and smoking status) are determined under non-parametric statistics (Mann–Whitney U-test (MWU) or Spearman test, adjusted for multiple testing using the Benjamini–Hochberg method). In the second step, for each feature significantly (FDR < 0.1) associated with defined phenotype status, it is checked whether it has significant associations with any potential confounder. If not, it is considered trivially unconfounded (not confounded (NC)). If at least one covariate also has significant association with the feature, then, for each such covariate, a post hoc test for confounding is applied. This test takes the form of nested linear model comparisons (likelihood ratio test for P values), where the dependent variable is the feature (X), and the independent variables are the QASD group (A) and the tested covariate (B) versus a model containing only the covariate (B), thus testing whether QASD status (A) adds explanatory statistical power beyond the covariate (B). If this holds (likelihood ratio test (LRT) P < 0.05) and the confidence interval for predictive inference in the LRT test does not include zero for all covariates (B), then disease status is strictly deconfounded (SD) concerning its effect on feature X; it cannot be reduced to any confounding factor. For each covariate (B) where significance is lost, a complementary modeling test is performed of the complementary model pairs, predicting (X) as a function of (A) and (B) versus a model containing (A) alone, thus testing whether the covariate (B) in turn is equally reducible to (A). If for at least one such covariate (B), (B) has independent effect (LRT P < 0.05) on top of (A), then the feature (X) is considered confounded by (B). However, if in none of the pairwise tests the original significance holds, then (A) and (B) are considered so correlated that their relative influence cannot be disentangled. We consider these cases ambiguously deconfounded (AD), in the sense that, for these cases, clear confounding influence can neither be concluded nor ruled out. The R package was applied to the present dataset considering medication status (statin, metformin and PPI intake, all as binary variables) and Metacardis clinical group as covariates and with country of recruitmet as random effect, and was runned on metagenomic (MGS, mOTU, Gut Metabolic Modules (GMM)) and metabolomic features (serum, urine) presents in at least 20% of the samples. For metagenomic features, abundances were corrected for bacterial cell count by multiplying by an index factor calculated as the bacterial cell count of the sample divided by the mean value of this bacterial cell count over the dataset as a whole ^71^. Our deconfounding pipeline considers linear effects related to drug categories. Still, we were not able to control for every possibly lifestyle confounding factor, making a lack of full confounding adjustment a limitation of our study.

### Statistical analyses and machine learning models

Data-management and statistical analysis were conducted using SAS version 9.3 and R (R statistical software, Vienna, Austria). The level of significance used was *P*<0.05, FDR<0.01, P for interaction<0.1.

Throughout the analyses, several adjustment strategies were used. First, for the analyzes concerning GMGR, a normally distributed variable, we systematically carried out and tested sensitivity analyses, taking into account the following confounding factors: age, sex, recruitment center, taking antibiotics, metformin and statins, BMI, which all are potential confounders identified in previous analyses from the MetaCardis consortium ^2,3,29,71^. Then, in accordance with what is done for dietary intake, models were adjusted on energy in kcal and aHEI score when we wanted to isolate the independent effects of nutritional quality. Finally, for specific omics features, we used the analysis pipeline published and developed in the MetaCardis study, thus allowing a better comparison between studies in the literature (see previous Drug deconfounding analysis section).

Predomics R package version 1.01^100^ was used to build interpretable predictive models of QASD status (high vs. medium, high vs. low, medium vs. low; High=QASD 1,2,3; Medium = 4; High= 5,6,7) based on MGS and metabolomic markers retrieved in the drug deconfounding analyses. Models were trained on three different algorithms including Random Forest as state-of-the-art (SOTA) method, and Binary and Ternary native predomics models describing simple ecological relationships in microbial ecosystems that were learned with Terbeam heuristics. Details of the different heuristics and BTR models can be found in Prifti et-al^100^. Models were trained on 80% of the data and evaluated for accuracy and AUC on a 10 times 10-fold cross-validation schema, and the results of the best algorithm were further explored to extract a family of best models (FBM), described as models whose accuracy is within a given window of the best model’s accuracy. This window is defined by computing a significance threshold assuming that accuracy follows a binomial distribution (p<0.05). No hyperparameter optimization was performed. Features included in the FBM were further explored in terms of prevalence across models and feature importance, described as the mean decrease accuracy (MDA) of the model after feature removal. The best model in each prediction task was evaluated on the 20% of the data excluded during the training process (hold-out data).

### Mediation analysis

Mediation analyses was used to infer causal mediation effects between MGS, serum metabolites and pairwise levels of the QASD score (High vs. Low, High vs. Medium, Medium vs. Low; Low=QASD 1,2,3; Medium = 4; High=5,6,7). To reduce the testing numbers, we focused these analyses on the MGS and metabolites with *OK_sd* status in the drug deconfounding pipeline and showing an absolute effect size greater than 0.1 on Cliff’s Delta between pairwise levels of the QASD score (features included in the Figures 3,4). In a first step, pairwise associations between metabolites and MGS were individually tested with linear regression analyses adjusted by country of origin, age, gender and intake of metformin, statin and protein pump inhibitors, and only significant pairs were retained for mediation analyses (FDR<0.05). MGS abundances were transformed with empirical normal quantile transformation method to ensure normality before the analyses. In the second step, similar regression framework with same adjustments was adopted to identify significant mediations under two possible hypothesis in terms of direction of the mediation using the *mediate* function of *mediation* V4.5.0 R package^101^. The first hypothesis delved into the role of MGS as mediators in shaping the relationship between the QASD score and the host’s metabolomic profile (Direction 1 in Figure 6A diagram). The second hypothesis aimed to explore whether the interplay between the QASD score and the host’s metabolome (mediator) could, in turn, influence the metagenomic landscape (Direction 2 in Figure 6A diagram). Significant mediations were considered those with FDR<0.05 for Average Causal Mediation Effect (ACME), Average Direct Effect (ADE) and Total effect. Similar analyses were carried out to infer causal mediation effects between levels of the QASD score, the microbial gene richness and the clinical status of the individuals in terms of corpulence (BMI) and metabolic status (glycated hemoglobin, HOMA-IR).

---

**Suppl. Figure 1:**
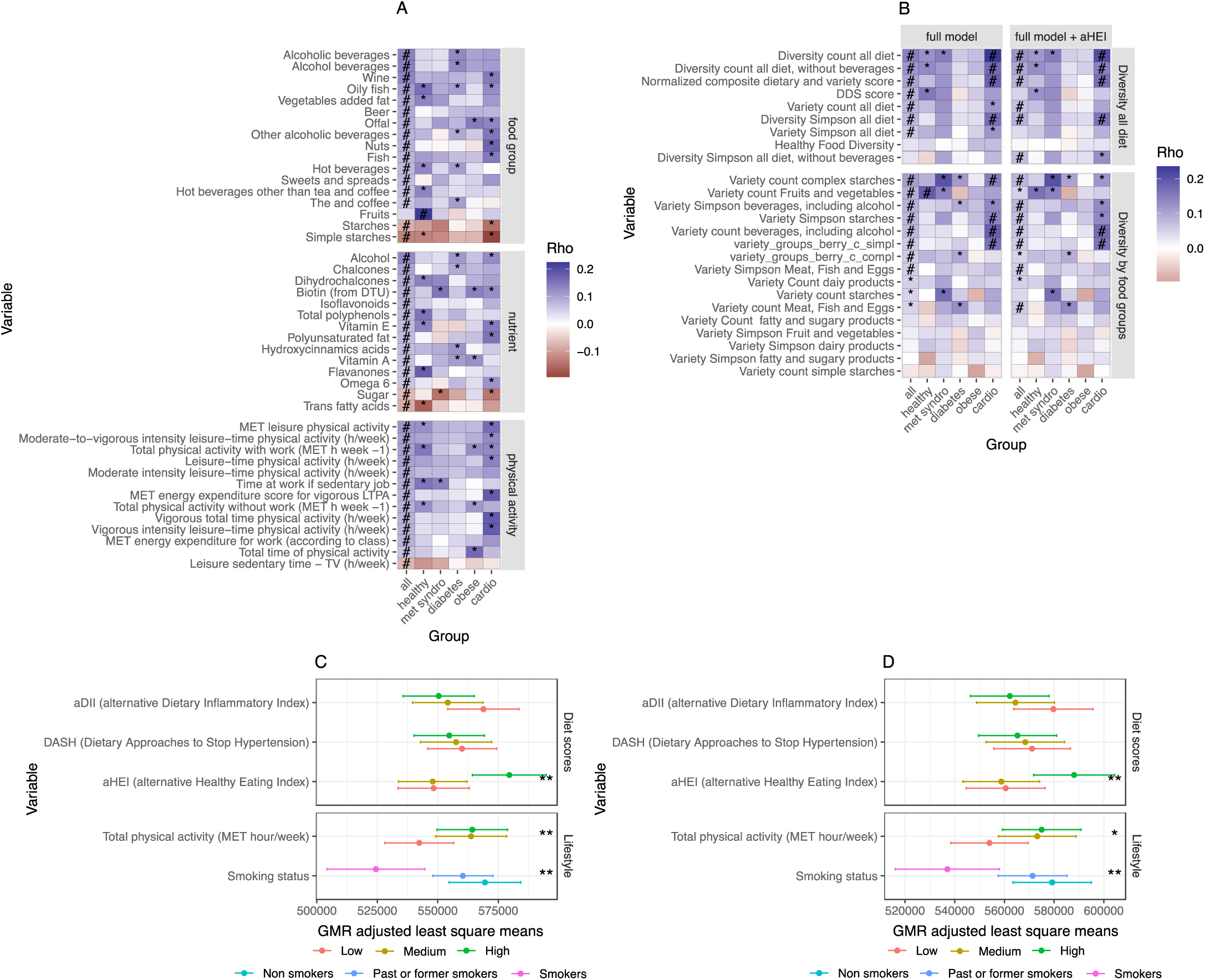
Sensitivity analysis on dietary and physical activity factors, diet variety and diversity scores associated with GMGR in MetaCardis population. (A) Heatmap representing the coefficients of partial Spearman correlations between GMR and nutritional and lifestyle factors (y-axis) in the full Metacardis cohort (all level in x-axis, corresponding to results shown in Figure 1A) and across individual clinical subgroups (additional x-axis levels). (B) Heatmap representing the coeffcients of partial Spearman correlations between GMR and dietary indexes (y-axis) in the full Metacardis cohort (all level in x-axis, corresponding to results shown in Figure 1C) and across clinical subgroups (additional x-axis levels). #=FDR<0.05; *=P-value<0.05, FDR>0.05; Partial Spearman correlations adjusted on age, center of recruitment, energy intake in kcal, antibiotics treatments (number of treatments in past five years), metformin, statin and PPI intake. (C) Estimated marginal means of GMGR across tertile levels of aDII, DASH, aHEI and Total physical activity and across levels of smoking status variable product of linear regression models adjusted on age, center of recruitment, energy intake in kcal, antibiotics treatments (number of treatments in past five years), metformin (yes/no), statin (yes/no) and PPI medications and BMI. (D) same as C additionally adjusted by clinical group. **=FDR<0.05; *=P-value<0.05, FDR>0.05, ANOVA tests on linear regression models.

**Suppl. Figure 2:**
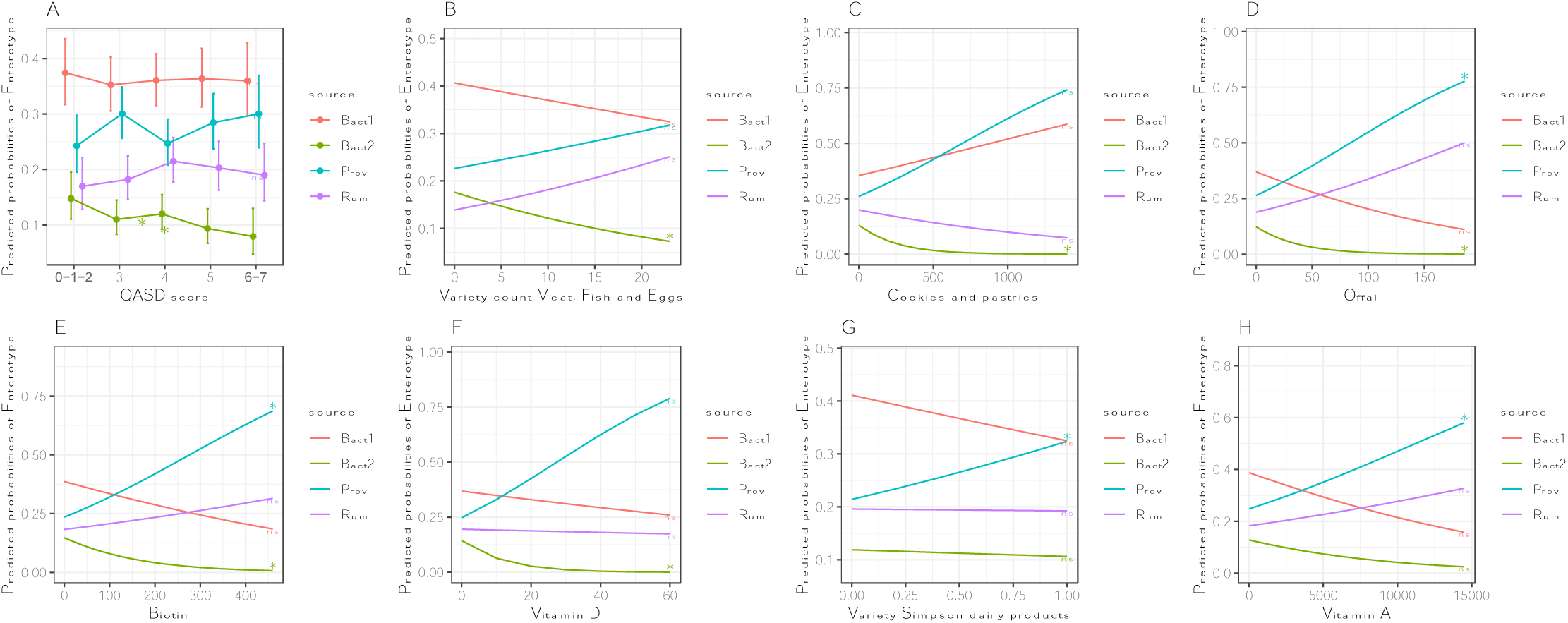
Nutritional variables showing significant associations with enterotype status of the Metacardis cohort: Logistic regression models have been fitted for each enterotype (Bacteroides 2, Bacteroides 1, Ruminoccocus, Prevotella) vs. 47 nutritional variables product of dbRDA analyses adjusted by age, gender, BMI center of recruitment, metformin, statin, and PPI intake. 8 variables showed significant associations with at least one enterotype status (*=p-value<0.05), which are represented in the form of predicted probabilities of each enterotype based on the corresponding logistic regression model. (A) High levels of the QASD score (5, and 6-7 levels) were associated to a significantly lower probability of Bact2 enterotype. In addition, Bact2 status is negatively associated with 5 additional nutritional variables (panels B-F) including “Variety count Meat, Fish and Eggs”, intake of different vitamins (biotin, Vitamin D), intake of “Cookies and pastries” ad Offal intake (high values of these variables significantly associated to low probability of being in Bact2 enterotype). Prevotella status is positively associated with intake of dairy products (Variety Simpson dairy products) and vitamin A (panels G,H), as well as with the consumption of offal (panel D) and biotin (panel E) in parallel with decrease of Bact2 status probability.

**Suppl. Figure 3:**
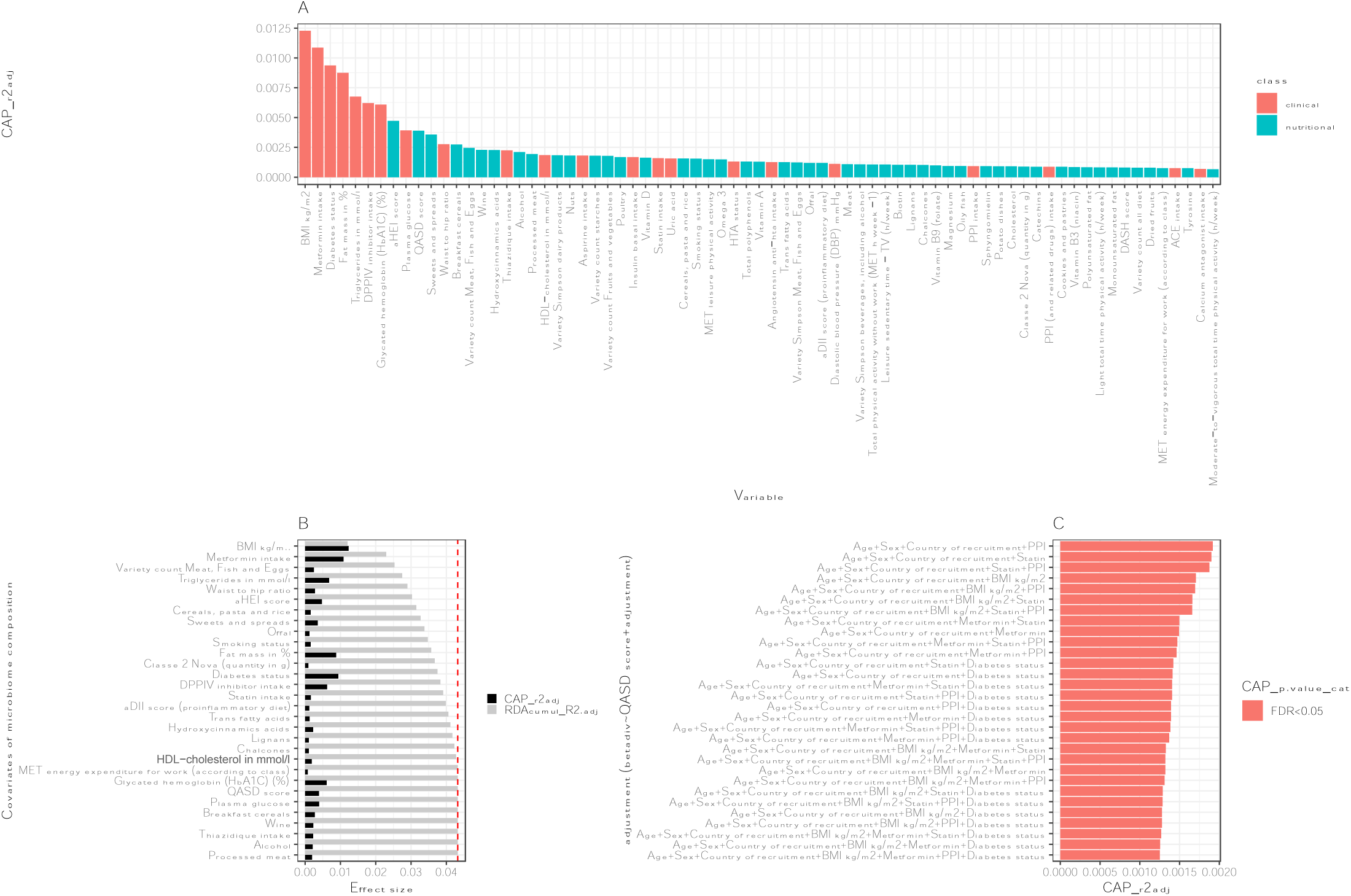
Evaluation of nutritional and bioclinical variables over microbiome composition in Metacardis cohort: (A) Barplot of effect sizes of 73 covariates (nutritional; clinical) with significant impact over microbiome composition in individuals of the MetaCardis cohort (n=1643; distance-based redundancy analysis, dbRDA; Bray-Curtis dissimilarity from MGS abundance data; FDR<0.01). (B) 30 of the 73 clinical and nutritional covariates with highest significant impact over microbiome composition in individuals of the Metacardis cohort (n=1643; distance-based redundancy analysis, dbRDA; Bray-Curtis dissimilarity from MGS abundance data), either independently (univariate effect sizes in black; FDR<0.01 in dbRDA) or in a multivariate model (cumulative effect sizes in gray). The cut-off for a significant non-redundant contribution to the multivariate model is represented by the red line (p-value<0.05 in stepwise model building). Full results in Supplemental Table S6 (C) Barplot representing the effect sizes of the impact of QASD score vs. microbiome composition (n=1643; distance-based redundancy analysis, dbRDA; Bray-Curtis dissimilarity from MGS abundance data) under different adjustment frameworks (y-axis; all adjustments support significant impact of QASD score over the microbiome composition in the adjusted dbRDA framework at FDR level).

**Suppl. Figure 4:**
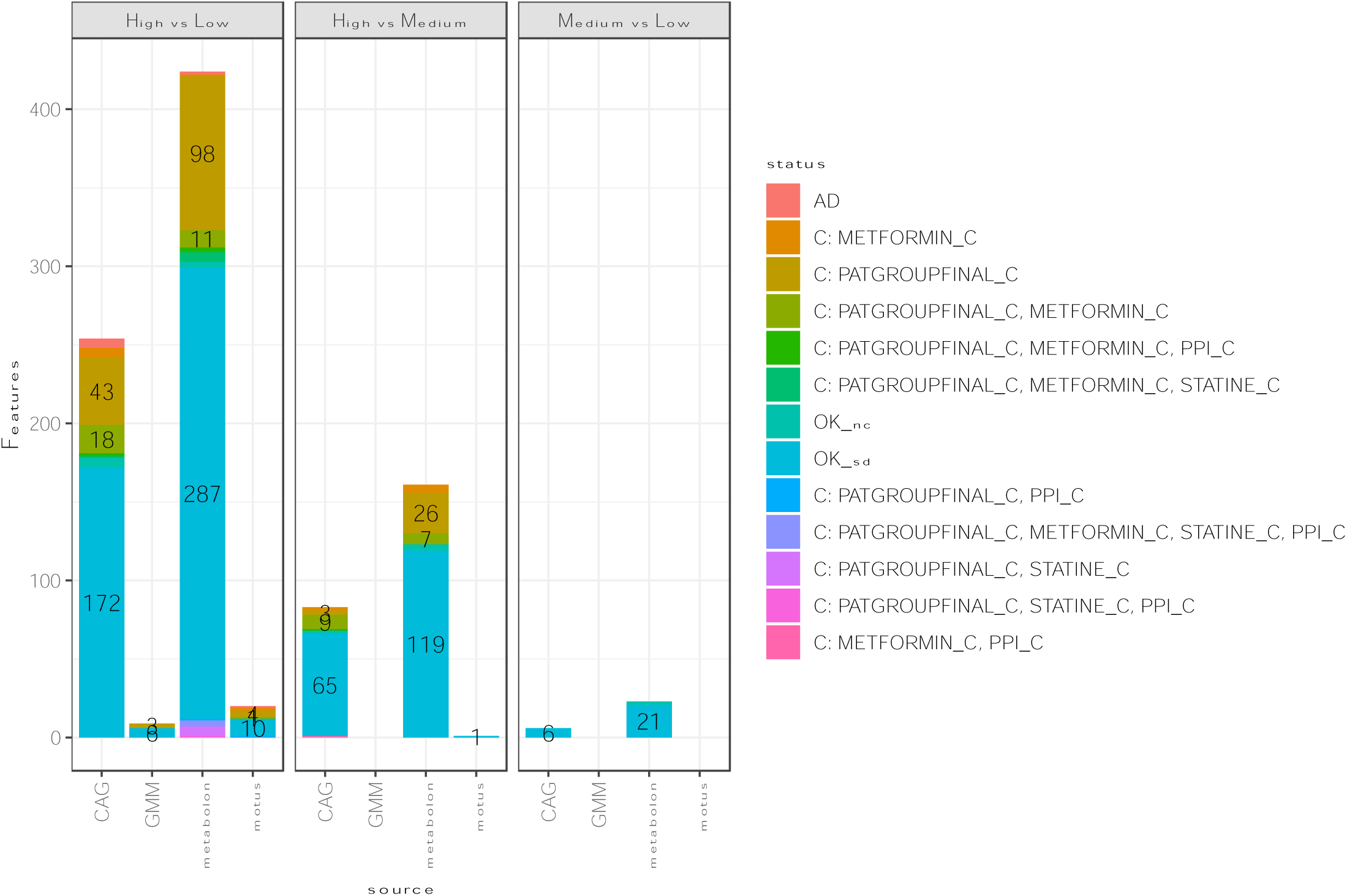
Summary of alterations in the microbiome and metabolome features (x-axis) between pairwise levels of the QASD score: Number of features in different spaces showing significant variations in between pairwise levels of the lifestyle score in metadeconfoundR analyses colored by the status of the association (AD: Ambiguously deconfounded, C: Confounded by the corresponding variables; OK_nc=No other covariates; OK_sd=Strictly deconfounded=The hardest label to reach, meaning that another covariate is associated to the feature but the signal can be reduced to the pairwise level of the lifestyle scores.)

**Suppl. Figure 5:**
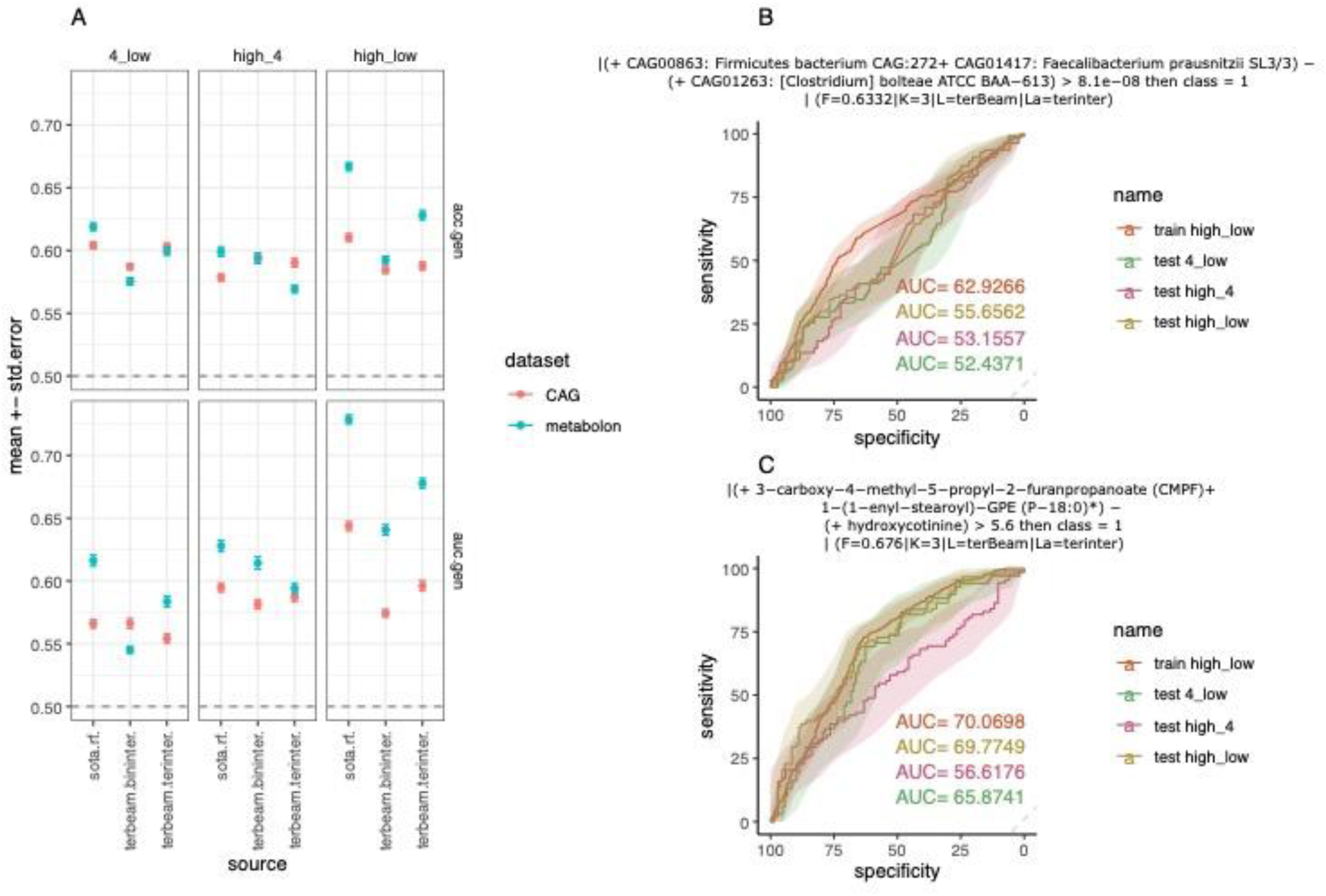
Predictive power of metagenomic and metabolomic features across pairwise levels of the QASD score. (A) mean ± standard error of accuracy (acc) and AUC of predictions between pairwise levels of the QASD score (3 levels decomposition) of best models based on random forest (sota.rf) and binary and ternary Predomics models built with terbeam learner. Models were trained on 80% of the data on 10 times 10-fold cross-validation schema on 135 MGS and 272 serum metabolites (metabolon quantification) product of deconfounding analyses (OK_sd status, FDR<0.1, absolute Cliff’s Delta effect size>0.1). (B) ROC curves of the best ternary model trained to classify individuals between high vs. low levels of the QASD score on the training data and holdout datasets (20% of the data not seen during training) based on MGS features. (C) Same as (B) based on metabolomic features

**Suppl. Figure 6:**
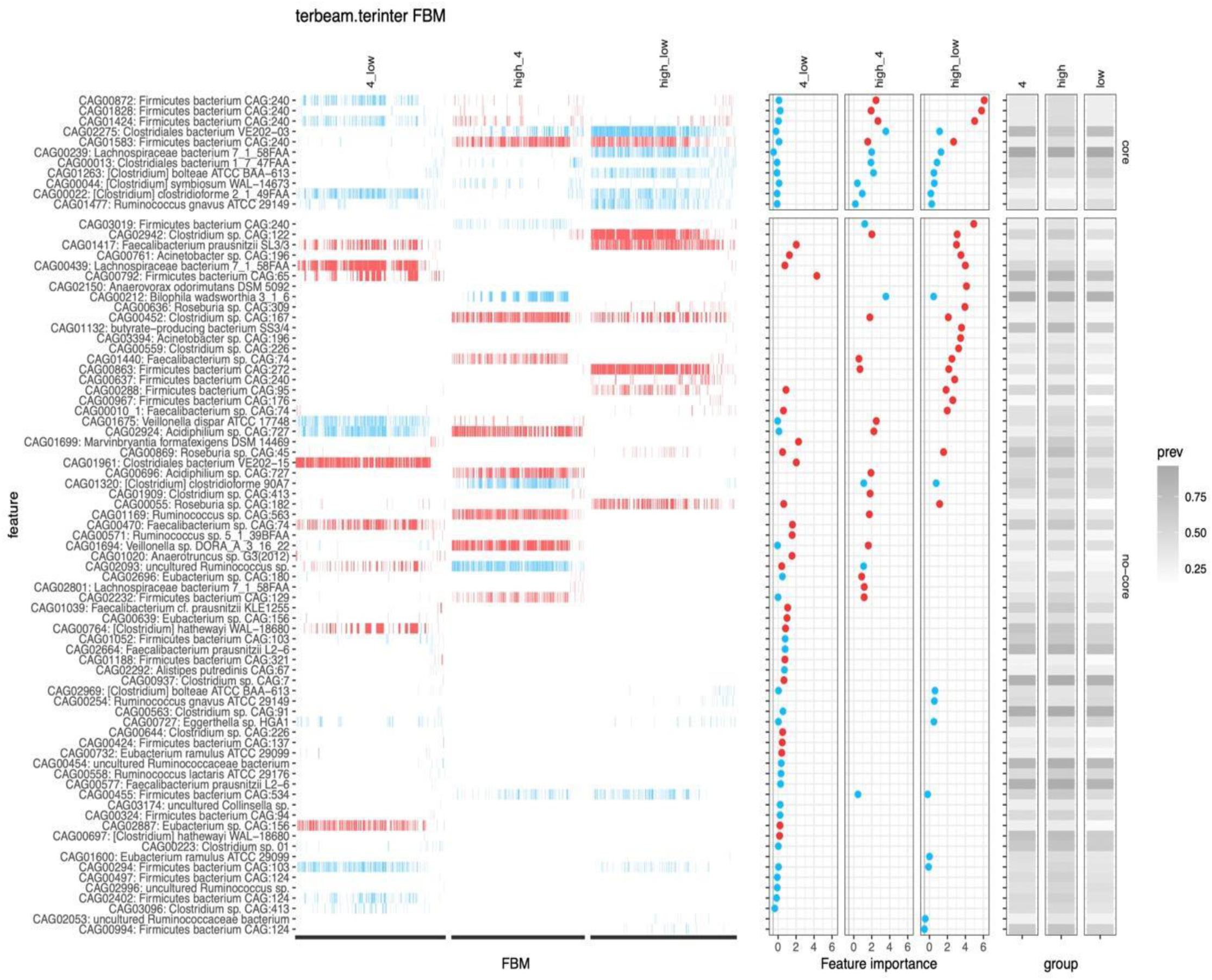
Summary plot of Predomics ternary models across pairwise levels of the QASD score based on MGS abundance profiles: Left panel represents the prevalence of 80 MGS retained across the Family of Best Models (FBM) of each binary prediction task (n=600, 797 and 875 models in 4 vs. low, high vs. 4 and high vs. low predictions). Middle panel represents the mean ± std. Error of feature importance variable (decrease accuracy when the feature is removed in the cross-validation process). The right panel represents the prevalence of the retained MGS across individuals in the 3 different levels of the QASD score. Blue color corresponds to MGS whose mean abundance is lower in the reference group in each pairwise comparison (4 in 4_low, high in high_4, high in high_low) whereas red color corresponds to MGS whose mean abundance is higher in the reference group in each pairwise comparison.

**Suppl. Figure 7:**
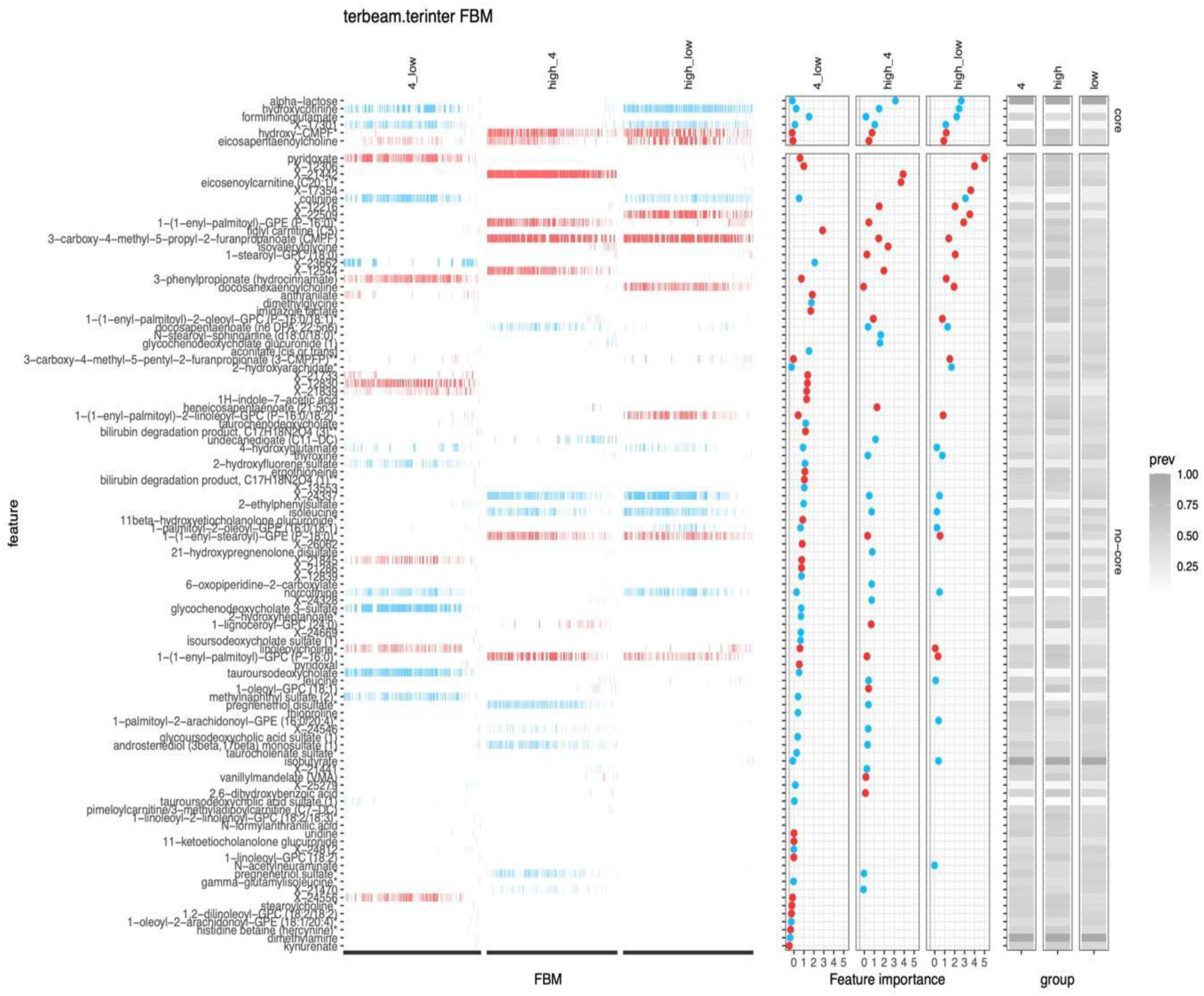
Summary plot of Predomics ternary models across pairwise levels of the QASD score based on metabolomics abundance profiles: Left panel represents the prevalence of 105 metabolites retained across the Family of Best Models (FBM) of each binary prediction task (n=847, 803 and 800 models in 4 vs. low, high vs. 4 and high vs. low predictions). Middle panel represents the mean ± std. Error of feature importance variable (decrease accuracy when the feature is removed in the cross-validation process). The right panel represents the prevalence of the retained metabolites across individuals in the 3 different levels of the QASD score. Blue color corresponds to metabolites whose mean abundance is lower in the reference group in each pairwise comparison (4 in 4_low, high in high_4, high in high_low) whereas red color corresponds to metabolites whose mean abundance is higher in the reference group in each pairwise comparison.

**Suppl. Figure 8:**
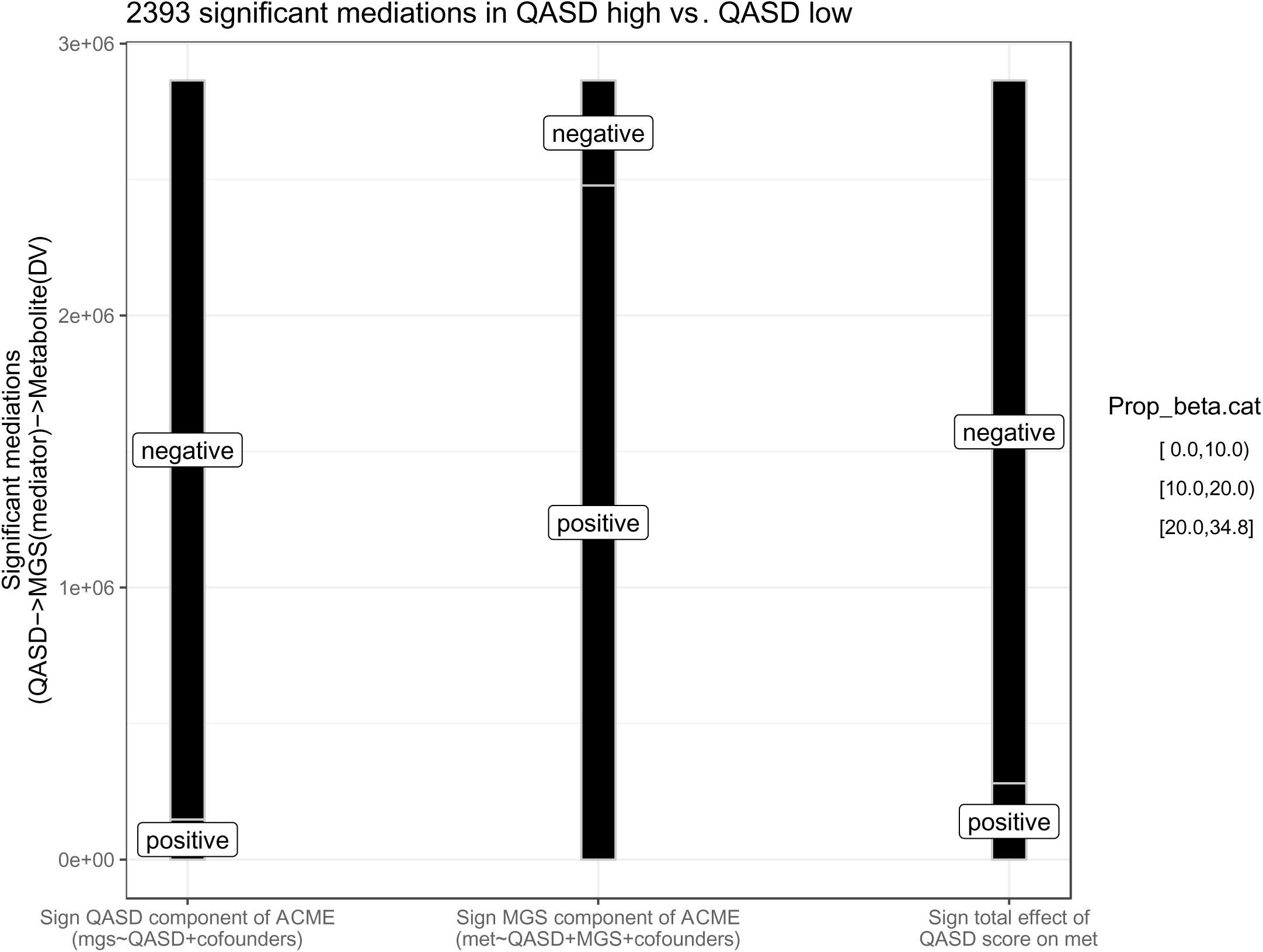
Summary of the different types of causal mediation relationships between QASD score (causal factor)->MGS (mediator)->Serum metabolite (outcome variable). Alluvial diagram showing the 2393 inferred causal mediation relationships in direction 1 (QASD score (high.vs. low) impacts on serum metabolite levels mediated by MGS abundances) decomposed by the sign of the beta coefficients in the QASD component of the ACME, the MGS component of the ACME and the total effect of the QASD score on the serum metabolite levels. Positive mediations are considered those where the sign of the MGS component of the ACME is positive (serum metabolite and MGS positively associated), whereas negative mediations are those where this sign is negative (serum metabolite and MGS negatively associated).

**Suppl. Figure 9:**
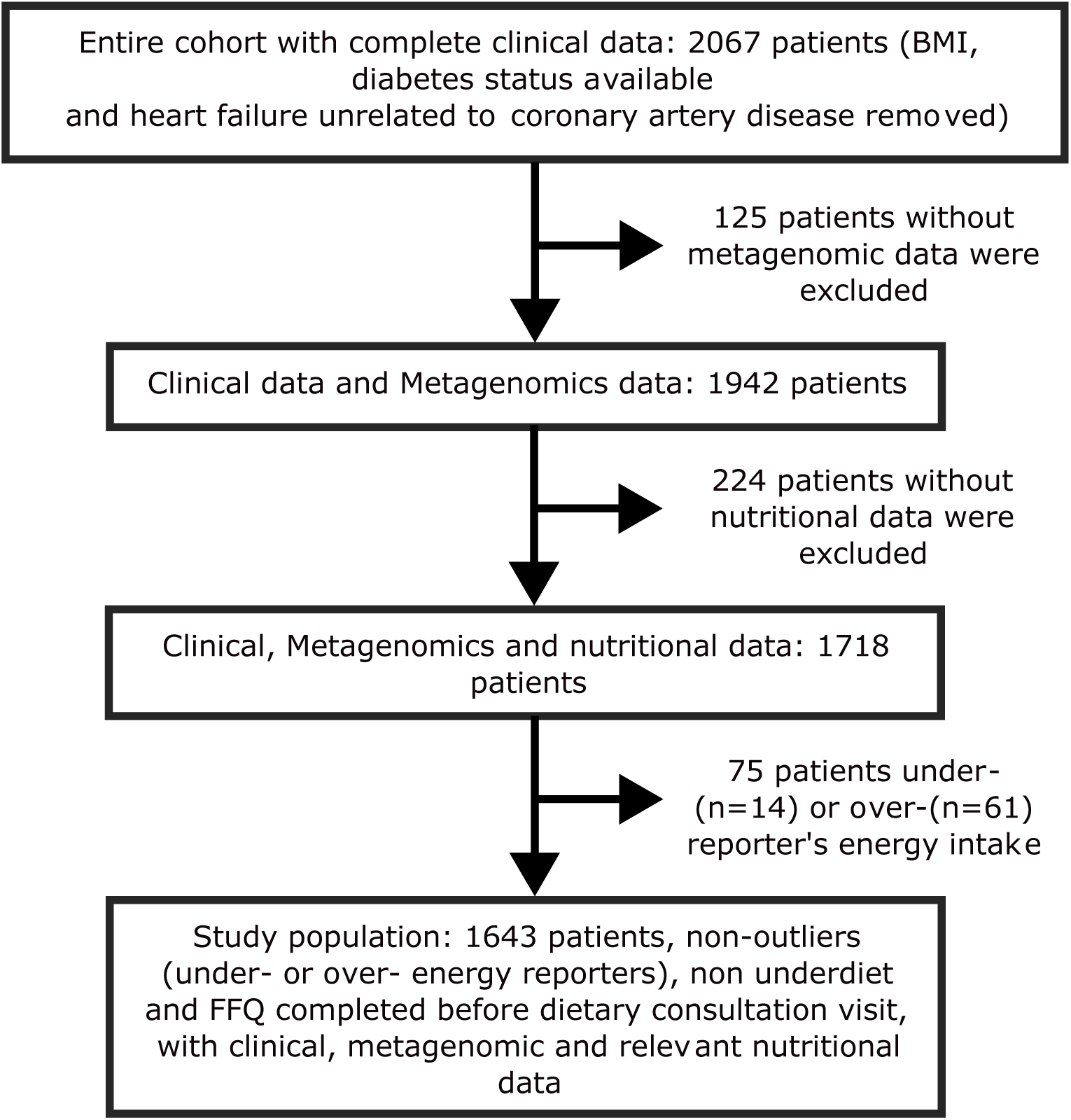
Flowchart of study design.

## Notes

### Competing Interest Statement

The authors have declared no competing interest.

### Clinical Protocols

https://clinicaltrials.gov/study/NCT02059538?cond=NCT02059538&term=NCT02059538&intr=NCT02059538&rank=1

### Author Declarations

The study protocol was approved by the Ethics Committee at the Medical Faculty at the University of Leipzig Germany (application number: 047-13-28012013) the ethical committees of the Capital Region of Denmark (H-3-2013-145) and the ethics committee Comite de Protection des Personnes (CPP) Ile-de-France III no.IDRCB2013-A00189-36 in France and was registered at https://clinicaltrials.gov/ (NCT02059538). The observational cohort design complied with all relevant ethical regulations, aligning with the Helsinki Declaration and in accordance with European privacy legislation. All participants provided written informed consent.

### Summary of Updates

Hello, the article was revised and replaces this version: MS ID#: MEDRXIV/2024/301195 Authors and text were corrected. Thanks

